# A simple computational model of population substance use

**DOI:** 10.1101/2022.09.11.22279817

**Authors:** Jacob T. Borodovsky

## Abstract

**Background:** Substance use behaviors and their etiologies are complex and often not amenable to traditional statistical analysis. Computational models are an increasingly popular alternative approach for investigating substance use. However, cumulative progress has been difficult because of a lack of standardization. This study aims to develop and evaluate a simple computational model that could serve as a common starting point for future computation-based investigations of substance use.

**Methods:** A two-state (“Using” a substance or “Not using” a substance) stochastic model with three manipulable parameters is used to reproduce the distributions of past 30-day alcohol, cannabis, and tobacco cigarette consumption frequencies (e.g., used on 5 days within the past 30 days) observed in the U.S. National Survey on Drug Use and Health (NSDUH) (years 2002-2019 combined). The model employs a path-dependent process: during each iteration (i.e., each “day”) of the simulation, each computational object chooses to use or not use a substance based on probabilities that are contingent on choices made in prior iterations. The Lempel-Ziv complexity measure was used to examine the resulting sequences of binary decisions (use ordon’t use) made by each computational object.

**Results:** The model accurately reproduces the population-level “U-shaped” distributions of past 30-day alcohol, cannabis, and cigarette use in the U.S. The path dependence function was required for reproducing these distributions. The model also suggests an “arc” of behavioral complexity stages: as the frequency of use increases, the complexity of decision sequences increases, peaks, and then decreases. However, decision sequence complexity still varied considerably among objects with similar frequencies of use.

**Conclusion:** A simple computational model that simulates individual-level sequences of substance use can reproduce the population-level distributions of substance use observed in national survey data. The model also suggests that complexity measures are a potentially helpful tool for examining substance use behaviors.

## INTRODUCTION

Substance use behaviors are likely driven by complex, nonlinear processes. Yet researchers frequently employ statistical methods that yield limited insight into such processes.^1–3^ Increasing recognition of this issue is pushing substance use research into new conceptual and methodological territories^4–8^ – particularly towards computational methods.^6,7,9^

The term “computational method” is generally considered to encompass a wide variety of primarily machine learning and simulation techniques.^10–12^ These techniques are being used to study substance use at many levels – ranging from neuroscientific to epidemiological.^13–16^ Simulation is a type of computational method that encourages explicit operationalization of assumed mechanisms as part of a computer program; computer experiments can then be conducted to determine if and how such assumptions generate outcomes of interest.^3,7,14,17–21^

Researchers are embracing simulation models to generate insights about a range of substance-related topics (e.g., substance use laws and regulations, social networks, economics).^16,22–28^ However, these models are often highly complex and use many idiosyncratic parameters and assumptions to examine specific circumstances. Consequently, the growing subfield of substance use simulation will have difficulty collaborating and building cumulative progress. One potential remedy is to develop simple yet broadly applicable “building-block” models of substance use to facilitate communication, collaboration, and new ways of thinking about the dynamics of substance use behaviors.

The first step in building such models is to identify a commonly observed macro-level phenomenon to replicate. One candidate phenomenon is the U-shaped distribution frequently observed in population substance use data. For example, each year the National Survey on Drug Use and Health (NSDUH) asks a random cross-section of the U.S. population questions such as, “*During the past 30 days, on how many days did you use marijuana or hashish?*” and “*During the past 30 days, on how many days did you drink one or more drinks of an alcoholic beverage?*”. ^*29*^ The distribution of responses to these questions is almost invariably U-shaped, with many low-frequency consumers at one end, many daily consumers at the opposite end, and fewer in between.

The next step is identifying and testing a micro-level process that plausibly governs the observed macro-level phenomenon. One starting place for identifying a relevant micro-level process is the Behavioral Science literature. Behavioral Science has established that the consequences (positive and negative) of a behavior will alter the probability of re-engaging in that behavior.^30^ Humans and other organisms learn that certain behaviors (e.g., consuming an intoxicating substance) produce reinforcing consequences (e.g., pleasurable subjective feeling, social validation, etc.) and subsequently seek to repeat the behaviors that produced the reinforcing consequences.^31,32^ This aspect of human behavior closely parallels the concept of “path dependence” in complex systems.^33^ Path-dependent systems evolve as a function of their own history, meaning that prior events impact future events (e.g., positive feedback loop).^34^ To date, various instantiations of path-dependent processes have been used successfully in economic and neurobiological models of substance use behaviors.^14^

The two aims of the present study were to (1) determine whether a simple computational model built around the concept of path dependence could reproduce empirical distributions of past 30-day frequency of alcohol, cannabis, and cigarette use in the U.S. population; (2) examine the complexity of individual-level behavior patterns to identify testable implications of the model.

## MATERIALS AND METHODS

This study took a pattern-oriented modeling^35–37^ approach by comparing simulated and empirical distributions of past 30-day substance use frequency (e.g., used a substance on 5 out of the past 30 days). The model is outlined using components of Hammond’s PARTE guidelines^38^ and Grimm et al.’s ODD guidelines.^37^ The study was conducted using Python 3.9 and Stata 17.

### Model Overview

This model uses a two-state, path-dependent, discrete stochastic process to reproduce the distributions of past 30-day consumption frequencies of alcohol, cannabis, and tobacco cigarettes observed in the National Survey on Drug Use and Health (NSDUH). This model shares conceptual similarities with other paradigms (e.g., “Individual-based”, “Agent-based”, “Multi-state”, and “Markov Chain” models.^9,25,34,37,39,40^). Broadly speaking, the model is populated with multiple computational objects. At each iteration (i.e., each “day”) of the simulation, each object decides whether to use the substance (represented by the number 1) or not use the substance (represented by the number 0). The decision to use or not use the substance is affected by the object’s properties. Specifically, each object has a unique probability of transitioning from the “Not using” state to the “Using” state and a unique probability of transitioning from the “Using” state to the “Not using” state. The latter probability changes based on the object’s history of use in previous iterations. To generate different distributions of past 30-day substance use frequencies, the modeler changes the values of three parameters: (1) Maximum Risk Factors Effect, (2) Minimum Protective Factors Effect, (3) Reinforcing Effect (these parameters are discussed in greater detail in the section “Initialization, Global Variables, Modeler Input”).

### Model Components

#### Object Properties

##### *“****Using****”* ***state (U)***, *“****Not using****”* ***state (N), and transition probabilities (Figure 1 and Table 1)***

**Figure 1.**
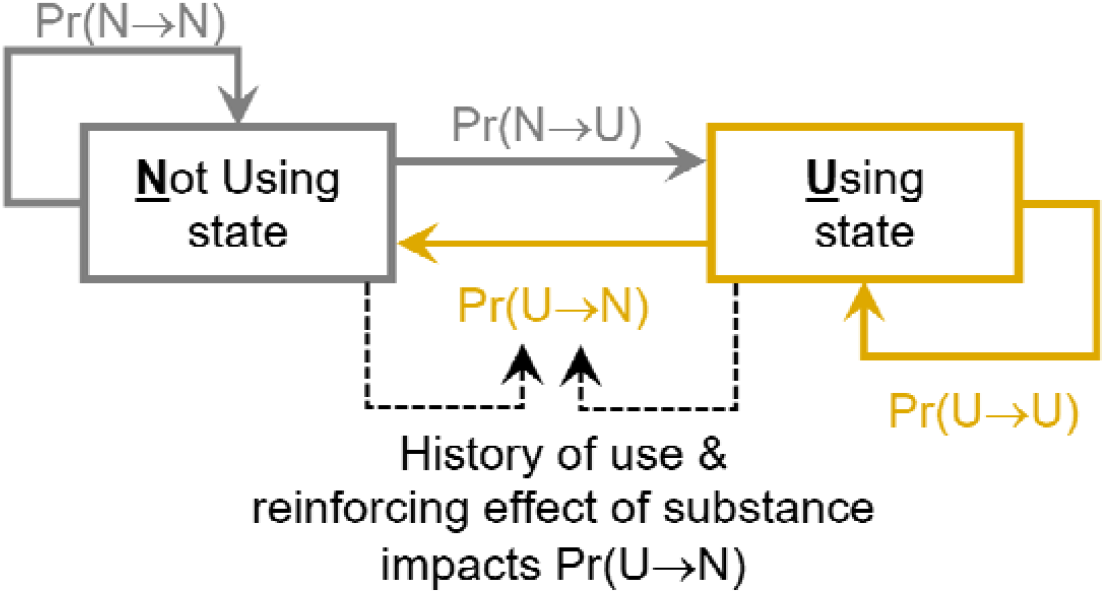
Conceptual structure of transition behaviors of an individual computational object Notes: (1) “Pr” =“Probability”; (2) “U” = “Using”; (3) “N” = “Not Using

**Table 1.** Model parameters

| Parameter | Description | Value is global or object-specific? | When is value calculated? | Calculation |
| --- | --- | --- | --- | --- |
| Maximum Risk Factors Effect | Maximum possible value of $\Pr(N \rightarrow U)$. | Global | iteration = 0 | Set by modeler. Value is a real number between 0 and 1 |
| Minimum Protective Factors Effect | Minimum possible value of $\Pr(U \rightarrow N)$ | Global | iteration = 0 | Set by modeler. Value is a real number between 0 and 1 |
| Reinforcing Effect | Theoretical value for a particular substance. | Global | iteration = 0 | Set by modeler. Value is a real number greater than zero. |
| $\Pr(N \rightarrow U)$ | Probability of transitioning from "Not using" state to "Using" state | Object-specific | iteration = 0 | Random draw from uniform distribution bounded between 0 and value of Maximum Risk Factors Effect |
| $\Pr(N \rightarrow N)$ | Probability of remaining in "Not using" state | Object-specific | iteration = 0 | $1 - \Pr(N \rightarrow U)$ |
| $\Pr(U \rightarrow N)$ | Series of probabilities of transitioning from "Using" state to "Not using" state. | Object-specific | iteration = 0<br>&<br>iteration > 0 | $\Pr(U \rightarrow N)_{i=0}$ is a random draw from uniform distribution bounded between value of Minimum Protective Factors Effect and 1.<br>$\Pr(U \rightarrow N)_{i>0} = \Pr(U \rightarrow N)_{i=0} * e^{(-\text{proportion days used} * \text{reinforcing effect})}$ |
| $\Pr(U \rightarrow U)$ | Probability of remaining in "Using" state | Object-specific | iteration = 0<br>&<br>iteration > 0 | $1 - \Pr(U \rightarrow N)$ |

Notes: (1) "i" = "iteration number"; (2) proportion of days used calculated using 30 as denominator; (3) value of $\Pr(U \rightarrow N)_{i>0}$ is re-calculated during each iteration that object is in the Using state

For each computational object, the simulation generates a sequence of binary states – “Not using” state (N) or “Using” state (U) – which represent the object’s pattern of substance use over time. The sequences reflect the object’s decisions to either remain in its current state or transition to a different state at each iteration for iterations i = 0,1,…n, where n is the total number of iterations in the simulation. Each object has a unique, fixed probability of transitioning from N to U called Pr(N⟶U), representing the object’s “Risk Factors Effect”. Pr(N⟶N) is the probability of remaining in N and is calculated as 1 − Pr(N⟶U). Each object also has a unique set of probabilities of transitioning from U to N called Pr(U⟶N), representing the object’s “Protective Factors Effect”. Pr(U⟶N) is initialized at i=0 and adjusted as the simulation progresses based on the object’s recent history of substance use. Finally, Pr(U⟶U) is the probability of remaining in U and is calculated as 1 − Pr(U⟶N). Additional details about these probabilities are provided in subsequent sections.

#### Object Action

##### Determining current state and deciding to use or not use

At the beginning of the current iteration, each object determines whether it is in the N state or the U state by checking whether or not it used the substance in the previous iteration. If the object did not use in the previous iteration, then it is in the N state at the beginning of the current iteration, and the probability that the object will use during the current iteration is the object-specific value of Pr(N⟶U). If the object used in the prior iteration, then it is in the U state at the beginning of the current iteration, and the probability that the object will *not* use during the current iteration is the object- and iteration-specific value of Pr(U⟶N). By the end of the current iteration, the object decides whether or not to use the substance during the current iteration. If the object decides to use, it records a 1 in its personal use history; if the object decides not to use, it records a 0 in its personal use history.

#### Object Action

##### Updating the value of Pr(U⟶N)

If the object uses during the current iteration, then the object’s unique probability Pr(U⟶N) is updated. The updated value – Pr(U⟶N)_i>0_ – is used by the object in the subsequent iteration when deciding whether or not to use. The value of Pr(U⟶N)_i>0_ is calculated as:

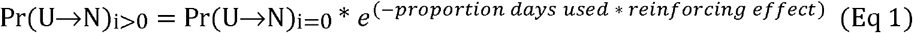

Where Pr(U⟶N)_i=0_ is a value established at initialization, “proportion days used” is a value between 0 and 1 calculated using the object’s personal substance use history results (containing either 0’s or 1’s) from the last 30 iterations. The Reinforcing Effect parameter is a global, fixed value used by all objects and is explained in greater detail below. Note that the same, fixed value of Pr(U⟶N)_i=0_ is always used to calculate each new value of Pr(U⟶N)_i>0_.

##### Time, Scheduling, and Environment

An iteration is completed after all computational objects have determined whether they use or not. Each iteration is considered to be equivalent to one day after the model has stabilized. Based on model stability testing results, 2000 iterations were used for generating results (see supplemental material). There is no spatial component to the model. Additionally, objects do not interact with each other directly or indirectly, which allows them to be statistically independent. This feature of the model mimics the statistical independence of NSDUH participants who are randomly sampled from across the U.S. and presumably do not know each other.

##### Initialization, Global Variables, Modeler Input

To generate different distributions of substance use, the modeler changes the values of three parameters: (1) Maximum Risk Factors Effect, (2) Minimum Protective Factors Effect, and (3) Reinforcing Effect. The Maximum Risk Factors Effect and Minimum Protective Factors Effect are set by the modeler as values between 0 and 1. Each object’s unique value of Pr(N⟶U) is determined at model initialization by a random draw from a uniform distribution with a lower bound set to zero and an upper bound set to the Maximum Risk Factors Effect. Similarly, each object’s unique value of Pr(U⟶N)_i=0_ is determined at model initialization by random draw from a uniform distribution with a lower bound set to the Minimum Protective Factors Effect and an upper bound set to one. The Reinforcing Effect parameter is a fixed value that applies to all objects (limitations underlying these concepts and assumptions are addressed in the discussion section).

#### Model Evaluation

##### National Survey on Drug Use and Health (NSDUH)

The National Survey on Drug Use and Health (NSDUH) is an annual, cross-sectional, multi-stage probability sample that assesses substance use behaviors among Americans age 12 and older with fixed household addresses.^41^ NSDUH data can be used to estimate the proportion of the U.S. population who have engaged in a particular substance use behavior. The present analyses used combined NSDUH data (years 2002 to 2019; N=1,005,421) to examine past 30-day frequency of alcohol, cannabis, and tobacco cigarette use. Specifically, for each substance, the proportion of past 30-day consumers who had used the substance on a given number of days was calculated and recorded (e.g., X% of past 30-day alcohol consumers had consumed alcohol on 9 days within the past 30 days) using the imputation-revised^41^ variables “iralcfm”, “irmjfm”, and “ircigfm” and appropriate survey weighting procedures.

Importantly, two modifications were made to the NSDUH data. First, the imputation-revised variables contained response values that were logically impossible for a discrete variable (e.g., used on 3.8 days). Therefore, values were rounded to the nearest integer. Second, empirical distributions of self-reported behaviors are often affected by the digit preference bias^42,43^ (e.g., tendency to estimate consumption in denominations of five). Therefore, respondents who reported using on 5, 10, 15, 20, and 25 days were randomly assigned with equal probabilities to either their original response value, one day greater, or one day less. For example, those who reported using on 15 days in the past 30 days were randomly re-assigned to either 14, 15, or 16 days.

##### Model stability

.Before the model could be used to simulate distributions of substance use, it was first necessary to determine the number of iterations required to obtain stable results. To determine the required number of iterations, simulated distributions were generated using different combinations of values of the Reinforcing Effect, Maximum Risk Factors Effect, and Minimum Protective Factors Effect parameters. During this process, the values of two of the three parameters were held constant while the value of the third parameter was varied. For example, distributions were generated using Reinforcing Effect parameter values of 3.0, 3.5, 4.0, 4.5, 5.0 while fixing the values of Maximum Risk Factors Effect and Minimum Protective Factors Effect to 0.2 and 0.7, respectively. Each combination of parameter values was tested in a simulation using 100,000 objects and 2000 iterations. For each simulation, the number of daily (i.e., 30/30 days) consumers was divided by the number of once-per-month consumers at the end of each 100-iteration interval (e.g., 400^th^ iteration, 500^th^ iteration, 600^th^ iteration, etc). A distribution was considered stable if this ratio was no longer changing substantially (examined qualitatively). Sensitivity tests using different starting seeds were conducted (supplemental material).

##### Effect of the path dependence function

To understand how the path dependence function (see Eq 1) impacts resulting distributions of past 30-day substance use, two simulations were conducted: one with the path function enabled, and one with the function disabled, with all other parameter values held equal across the two conditions.

##### Model calibration

For each substance (alcohol, cannabis, tobacco cigarettes), a least-squares approach was used to identify the best fit between a simulated distribution of past 30-day use proportions and the empirical (NSDUH) distribution of past 30-day use proportions. To sweep the parameter space, simulated distributions were generated using combinations of values within given ranges for three parameters: Maximum Risk Factors Effect, Minimum Protective Factors Effect, and Reinforcing Effect. More details about the calibration process can be found in the supplemental material.

##### Complexity of use patterns in different simulated populations

Each object in a simulation records its history of decisions to use or not use which is represented as a series of ones and zeros. For example, an object with a history of 000111 represents three consecutive decisions to not use followed by three consecutive decisions to use. The Lempel-Ziv algorithm^44^ was used to measure complexity of these use histories (Table 2). The mean number of Lempel-Ziv sequences was calculated for different simulated populations generated by different combinations of values of the three manipulable parameters (Reinforcing Effect, Maximum Risk Factors Effect, Minimum Protective Factors Effect).

**Table 2.** Example binary sequences of substance use and corresponding number of Lempel-Ziv sequences

| Substance use pattern<br>(0 = no use, 1 = use) | Number of Lempel-Ziv sequences |
| --- | --- |
| 0000000000 | 4 (less complex) |
| 1111111111 | 4 |
| 0101010101 | 5 |
| 0000111110 | 6 (more complex) |

## RESULTS

### Model Stability

Producing different distributions of substance use by varying the values of the Reinforcing Effect, Maximum Risk Factors Effect, and Minimum Protective Factors Effect parameters generally suggested that the ratio of 1/30 day users to 30/30 day users was stable before reaching 1000 iterations. Given this result, a conservative 2000 iterations was chosen for the model evaluation process.

### Effect of the path dependence function

Figure 2 displays simulated distributions in which the path dependence function (Eq1) is disabled (left side of Figure 2) and enabled (right side of Figure 2). The results provide evidence that enabling the path dependence function is responsible for generating the “U-shaped” distribution of interest.

**Figure 2.**
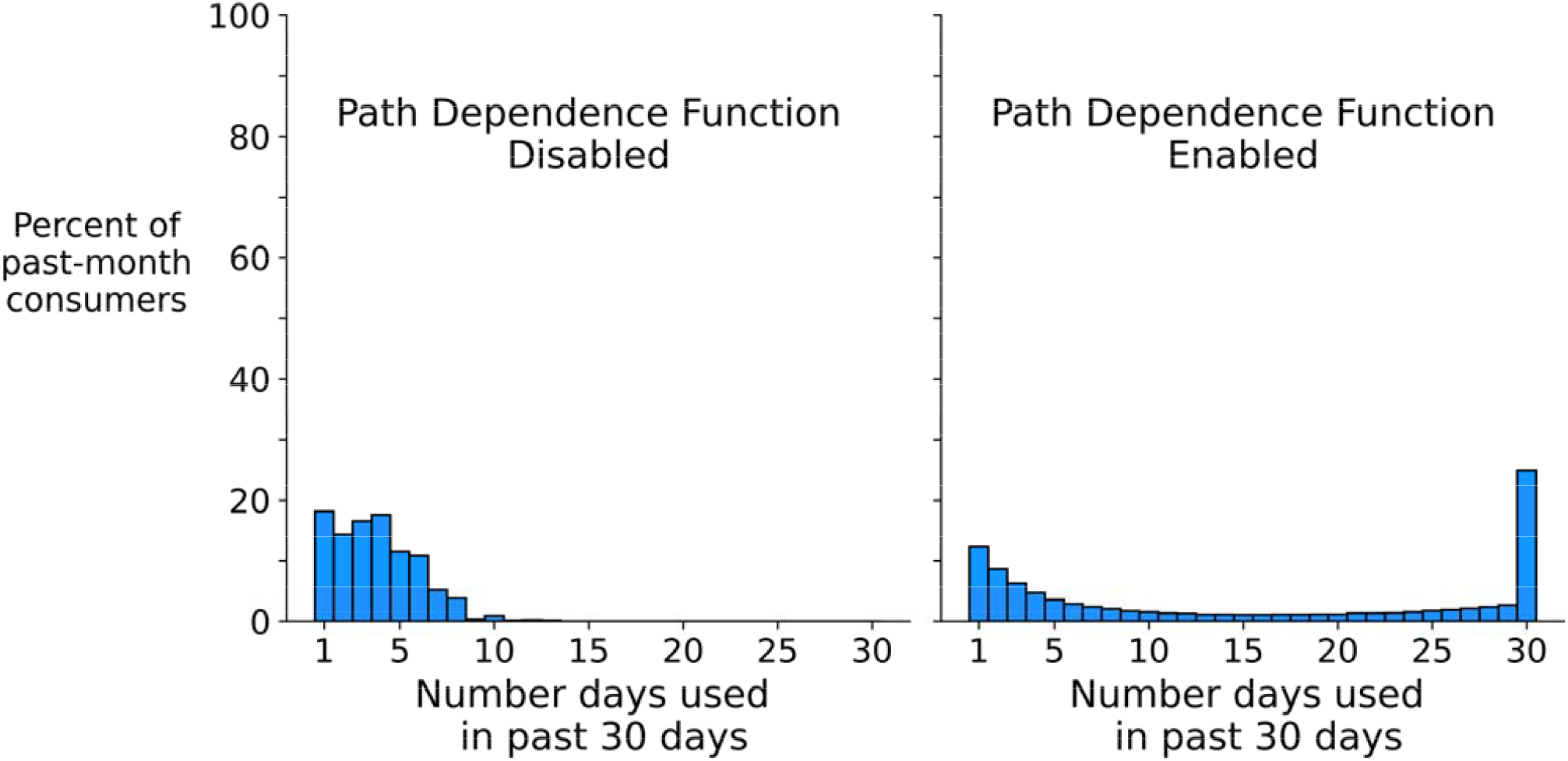
Simulated distributions of past 30-day substance use frequencies produced by disabling and enabling the path dependence function Note: 100,000 objects and 2000 iterations used to created simulated distributions

### Model Calibration

The right side of Figure 3 displays the simulated distributions produced by the least squares-optimized parameter values next to the corresponding distribution from the NSDUH data (left side of Figure 2). Note on the right side of the figure that a greater value of the Reinforcing Effect parameter (3.0 for alcohol, 3.8 for cannabis, 4.7 for tobacco cigarettes) corresponded to a greater proportion of daily (30/30 days) consumers. When comparing alcohol and cannabis, there is a similar increase in the optimized values of the Maximum Risk Factors Effect (alcohol=0.165 vs. cannabis=0.175) and the Minimum Protective Factors Effect (alcohol=0.5 vs. cannabis=0.58). However, this trend did not continue for cigarettes. The optimized values of Maximum Risk Factors Effect and Minimum Protective Factors Effect for cigarettes were lower than those of alcohol.

**Figure 3.**
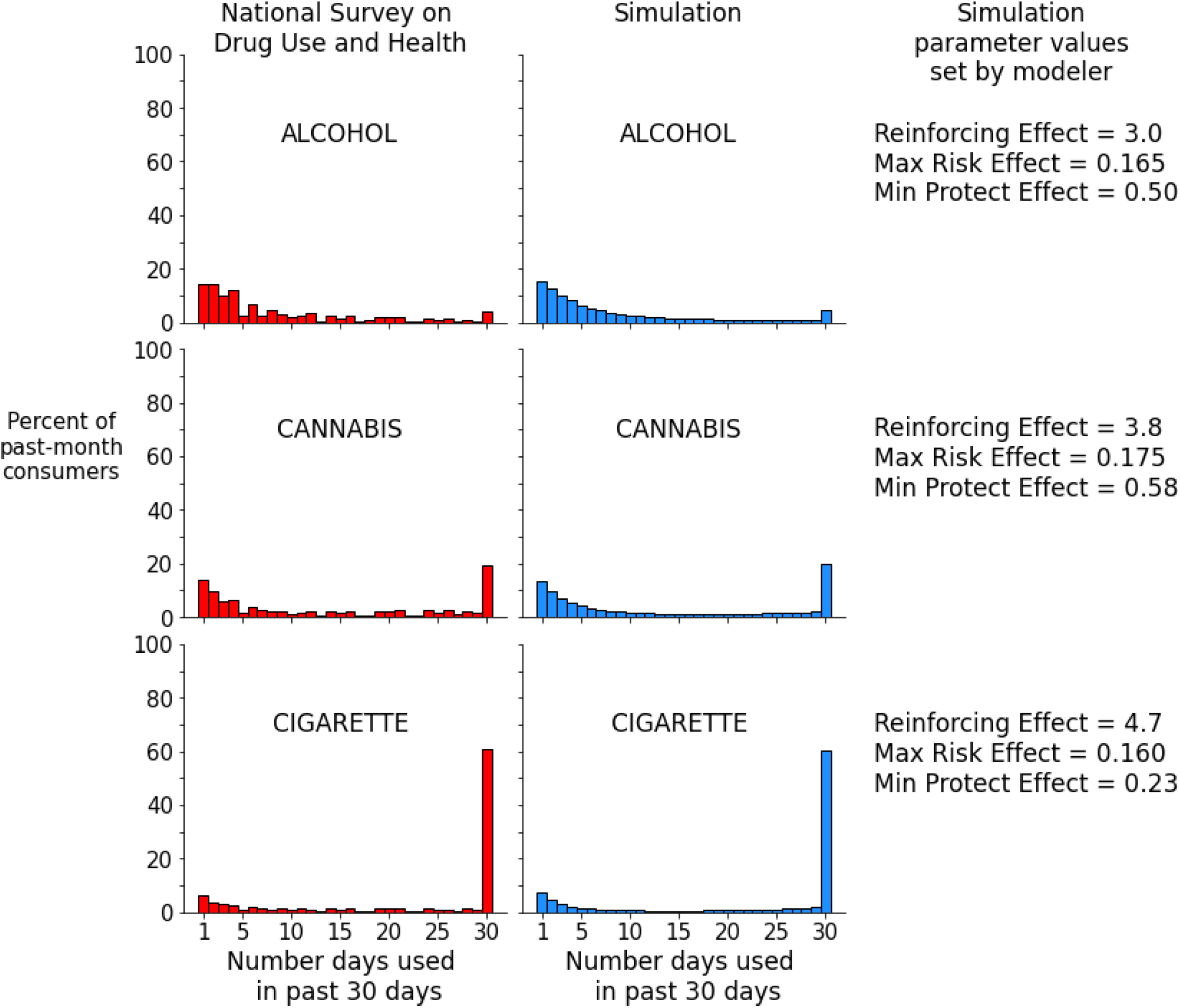
Survey-based distributions vs. simulated distributions of past 30-day substance use frequencies Note: 100,000 objects and 2000 iterations used to created simulated distributions

### Proportion of days used vs. complexity of historical use pattern (Figure 4 and Figure 5)

Figure 4 displays the mean complexity of past 30-iteration use patterns among objects. The objects were divided into three groups based on the ratio of total number of use days (i.e., total iterations in which the object recorded a 1) to total number of iterations in the simulation (2400). For example, the line with the circle markers in Figure 4 represents the changes in the mean past 30-iteration complexity of objects that were destined to use on ≥70% of all iterations (i.e., “lifetime high-frequency consumers”) in the simulation. The key dynamics to note are the changes in mean complexity over time and the value at which a subgroup’s complexity stabilizes. For example, lifetime high-frequency consumer objects exhibit a mean complexity trajectory that increases, decreases, and then stabilizes below that of the “lifetime moderate frequency consumers” subgroup (i.e., below the mean complexity of objects that used between 20-70% of iterations).

**Fiqure 4.**
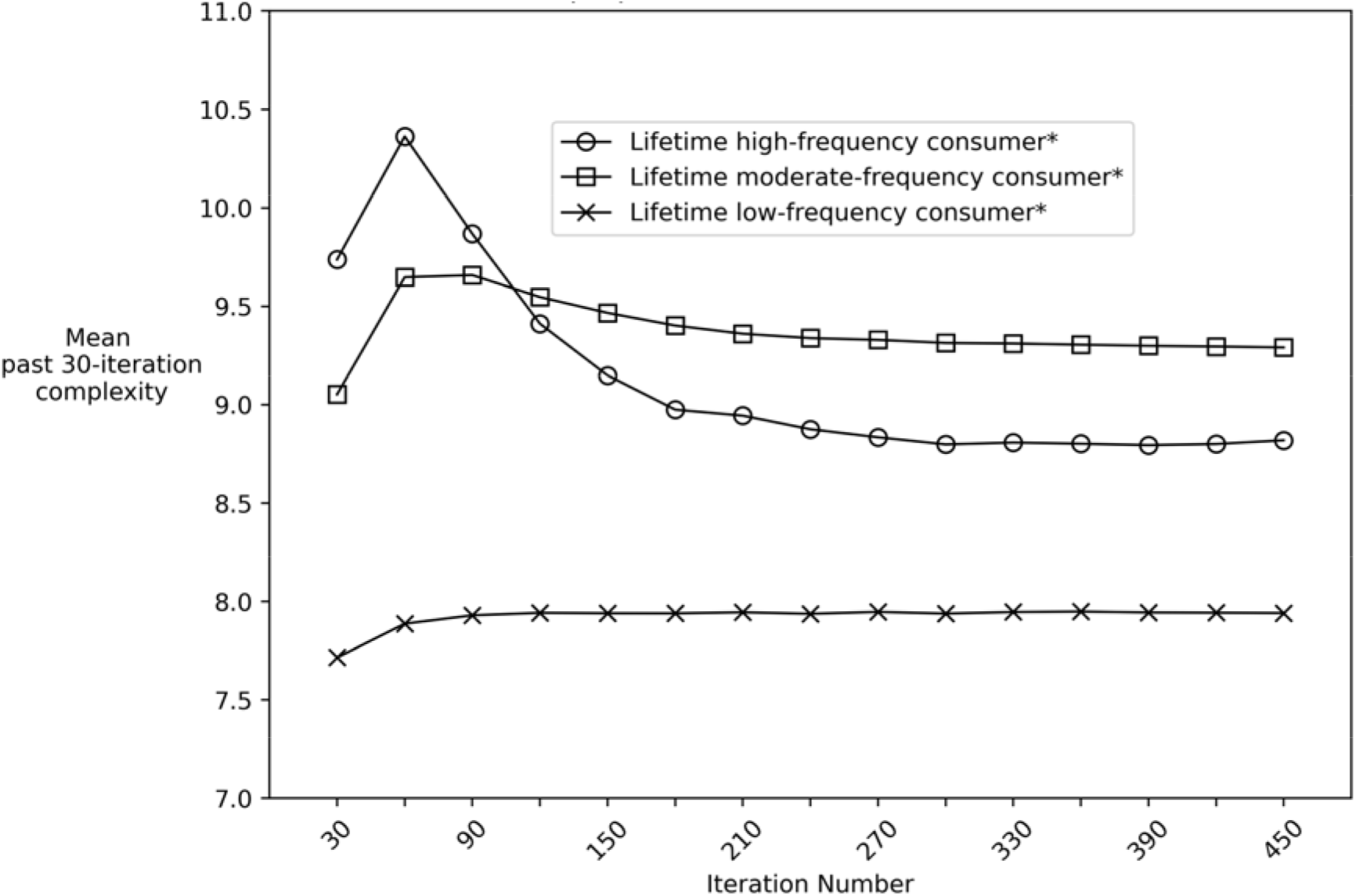
Mean complexity of 30-iteration intervals stratified by proportion of total iterations used Notes: Simulation conducted using 100.000 objects and 2400 iterations *High-frequency = used on a 70% of 2400 iterations *Moderate-frequency = used on >20% & <70% of 2400 iterations *Low-frequency = used on < 20% of 2400 iterations

However, Figure 5 demonstrates that even among objects with the same proportion of iterations used, there is still variability in complexity scores. Each of the four subgraphs in Figure 5 represents the entire 2000-iteration history of decisions to use and not use for four different computational objects.

**Figure 5.**
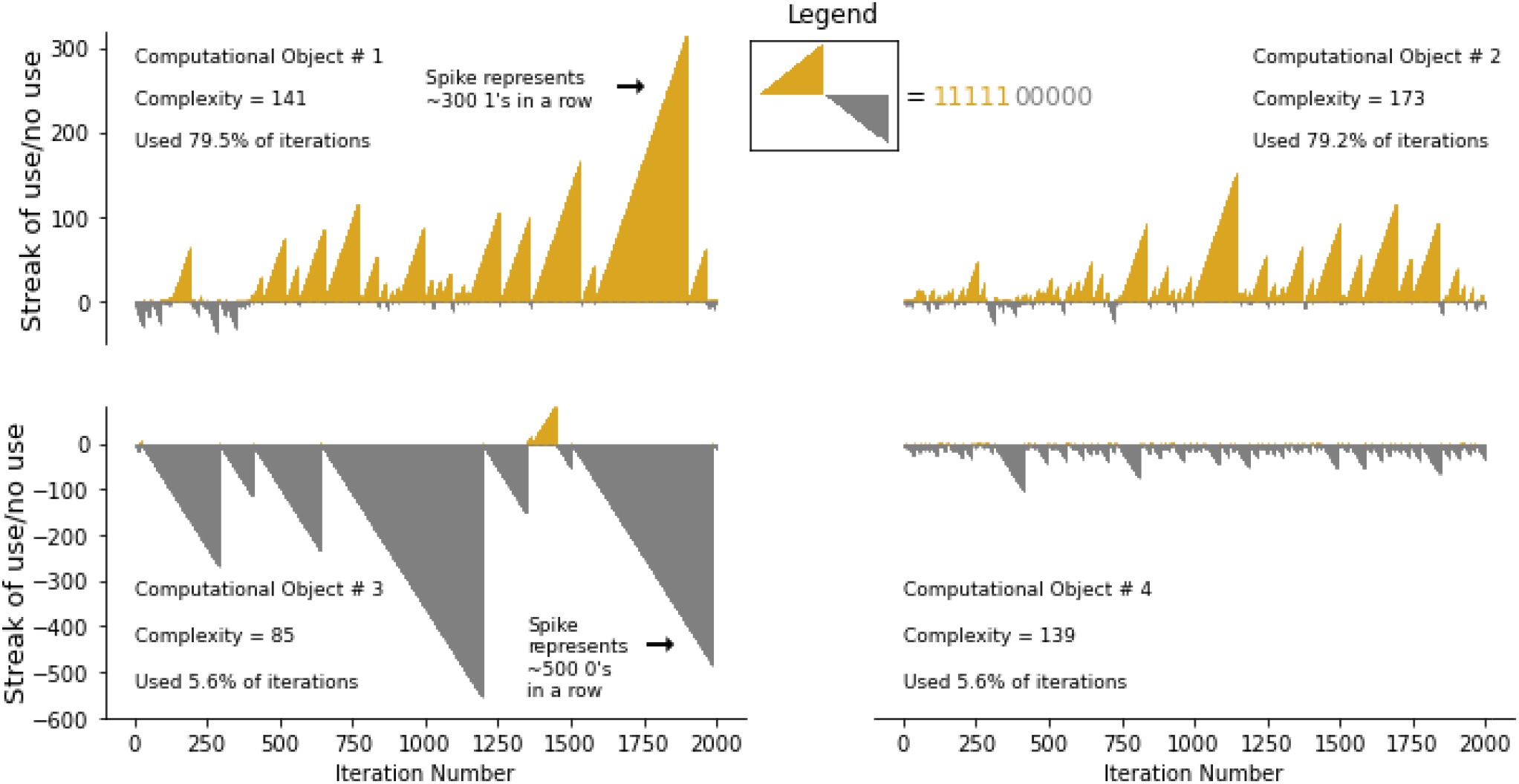
Pattern of Using (l’s) and Not Using (0’s) streaks over 2000 iterations for four different computational objects

Every gold “spike” is a cumulative count of a consecutive series of ones (i.e., consecutive series of decisions to use); every gray “spike” is a cumulative count of a consecutive series of zeros (i.e., consecutive series of decisions to not use). The central point conveyed by Figure 5 can be gleaned by comparing the use history of object one (top left subgraph) to the use history of object two (top right subgraph). Both objects used ∼79% of iterations, and yet the use pattern in the top left graph is less complex (L.Z. Complexity = 141) than the use pattern in the top right graph (L.Z. Complexity = 173). Comparing the object histories in the bottom left and bottom right subgraphs yields a similar conclusion. These results demonstrate that, in principle, the same proportion of days used (a common metric used for studying substance use behaviors) can yield different complexities of use.

## DISCUSSION

This study outlined a model that simulates individual-level sequences of substance use to reproduce population-level distributions of substance use observed in national survey data for three substances. The model uses a two-state, path-dependent, discrete stochastic process and requires only three inputs from the modeler. This model’s simplicity and flexibility could make it a useful “building block” for other computational models of substance use. This model could potentially also be modified to study various social, geographic, or economic dynamics that underpin substance use initiation, maintenance, and cessation.

The concept of path dependence plays a central mechanical and theoretical role in this model. From a mechanical perspective, path-dependent dynamics arise because an object’s current probability of a behavior was programmed to be contingent on the object’s prior behaviors. The results suggest that this mechanism is critical for producing the “U-shaped” distributions of population behaviors observed in the NSDUH data. From a theoretical perspective, the contingency between current and prior behavior represents the notion that consuming a “highly addictive” substance many times in the recent past is associated with a high probability of continuing to use that substance in the present. The plausibility of this conceptual leap is discussed in greater detail below. However, to drive the theoretical point further, consider the following hypothetical scenario. Imagine that a “maximally addictive” drug (i.e., maximally path-dependent drug) existed and that a person was guaranteed to become a daily consumer of this drug after trying it just once. Such a drug could only produce an essentially binary distribution of past 30-day consumption frequencies: X% of the population having never used the drug (i.e., 0/30 days of use) at one end of the distribution, and nearly 100-X% of the population with 30/30 days of use at the opposite end of the distribution (the exceptions in the middle of the distribution being those who began using the drug for the first time within the past 30 days). This example represents the technical (albeit highly unlikely) upper boundary of what we can expect empirical distributions of past 30-day substance use to look like in a population.

This study also used the Lempel-Ziv procedure to summarize the complexity of binary (Use or No Use) sequences of behavior. The simulated results suggest that several phenomena are, in principle, observable in empirical substance use data: (1) For a given combination of risk and protective factor distributions (i.e., a given combination of Pr(N⟶U) and Pr(U⟶N)), we expect that a peak mean complexity of population substance use histories exists at a particular reinforcing effect value (i.e., particular “addictiveness”); (2) Longitudinally, individuals who are destined to become daily consumers pass through three “phases” of consumption pattern complexity: low complexity pattern, high complexity pattern, and then low complexity pattern again; (3) Similarly, the cross-sectional complexity of substance use histories in a population may have a concave relationship with the cross-sectional frequency of substance use: on average, low-frequency and high-frequency consumers have lower-complexity histories of use, whereas moderate consumers have higher-complexity histories; (4) However, among objects with similar frequencies of use, the complexity of individual substance use histories can vary substantially (e.g., two individuals who both used on 10 days in the past 30 days can have different complexity scores). This last point raises an intriguing possibility: complexity measures (e.g., Lempel Ziv, Approximate Entropy, Permutation Entropy) may have clinical utility if they improve our ability to discriminate future outcomes among seemingly homogenous groups of substance consumers.

There are a variety of limitations, assumptions, and caveats to consider. This model was programmed so that the probability of current substance use is contingent on historical patterns of substance use because “*Substance use behaviors are an example of a psychological system that exhibits feedback: the use of substances and the circumstances in which it takes place can impact one*’*s life in both the short-term and long-term, creating environments that are reinforcing thereby impacting the amount of substance use*”^45^. However, this model oversimplifies the complex pharmacological, genetic, and environmental interactions that drive substance use behaviors in the real world.^46,47^ For example, the model employs a “Reinforcing Effect” parameter even though it is technically incorrect to say that a substance has intrinsic reinforcing “properties”, or that one substance (e.g., nicotine) is “more addictive” than another (e.g., alcohol).^31,48^ Nonetheless, the use of a reinforcing effect parameter seems justified given that, in aggregate, the conditional probability of developing a substance use disorder varies across pharmacologically distinct substances.^49,50^

An additional limitation concerns the two unique probabilities – Pr(N⟶U) and Pr(U⟶N)_i=0_ – assigned to each object in the model. These probabilities are meant to summarize the object’s unique combination of biopsychosocial risk and protective factors that drive the object’s binary decisions. This is an extremely strong assumption, and many valid arguments could be made for alternative conceptual frameworks. However, the approach taken in this study is not entirely out of step with modern research practice. Researchers routinely assign individual outcome probabilities by modeling linear combinations of relevant risk and protective factors in statistical models. Furthermore, there is a long precedent of using stochastic and state-based models to model the probability of binary outcomes.^51,52^ A related issue is that the model also assumes Pr(N⟶U) and Pr(U⟶N)_i=0_ are uniformly distributed in the population. We conducted sensitivity tests using non-uniform beta distributions and found that the model produces the same results and only requires different initialization values for the three modifiable parameters (Maximum Risk Factors Effect, Minimum Protective Factors Effect, Reinforcing Effect; see supplemental material). Therefore, it may be more important to determine the interpretation of the Pr(N⟶U) and Pr(U⟶N)_i=0_ probabilities rather than the distribution from which these probabilities are drawn.

Limitations concerning the concept of time in this model also warrant discussion. This simulation was designed to produce longitudinal data that could be analyzed cross-sectionally and compared to cross-sectional NSDUH data. In this study, one iteration of a simulation was considered equivalent to one day in the post-stabilization phase of the simulation (i.e., after the first 1000-2000 iterations). However, during the pre-stabilization phase of the simulation, the unit of time represented by an iteration is less clear. For example, in the real world, some individuals may rapidly escalate to daily use within three months of initiation; others may vacillate over several years with periods of infrequent use and periods of frequent use before finally settling into a stable pattern of daily use. Objects in the simulation also exhibited various use trajectory shapes during the pre-stabilization phase. However, because of a lack of empirical data, it is unknown whether treating each pre-stabilization iteration as a single day would make the simulated trajectories temporally consistent with real-world trajectories.

It is also important to consider that different models can reproduce the same data^53^, and it may be difficult to discern which models exhibit the greatest fidelity to real-world mechanisms and processes. Moving forward, it is essential that theoretically-oriented models such as the one presented here remain tethered to reality through ongoing comparison to empirical data.^47,54,55^ Overall, simulation models of substance use, such as the one presented here, are perhaps best viewed as one of several complementary epidemiological approaches that can be used to triangulate answers to questions of interest.^2,7,56,57^

## Conclusions

In sum, this study illustrates that a simple computational model can be used to accurately approximate empirical distributions of population substance use. Incorporating path dependence functions and complexity measures into simulation models could generate new insights into the links between individual- and population-level patterns of substance use, as well as produce results with testable clinical and public health implications.

## Supporting information

Model Code

## Data Availability

Python code used to produce simulated data are provided as supplemental material. NSDUH data are available online https://www.datafiles.samhsa.gov/dataset/nsduh-2002-2019-ds0001-nsduh-2002-2019-ds0001

## Acknowledgements

The author would like to acknowledge the following individuals: Ross Hammond, Douglas Luke, Matthew Kasman, Robert Purcell, Joe Ornstein, Mohammad Habib, Leah Koenig.

