## Supplemental Material for "A simple computational model of population substance use"

**SUPPLEMENTAL MATERIALS**

**Model Stability**

Results from model stability testing are provided in the figure below. The three subgraphs correspond to testing different values of the three modifiable parameters: Reinforcing Effect, Maximum Risk Factors Effect, and Minimum Protective Factors Effect. In each subgraph, the values of one parameter are varied while the values of the other two are held constant. The outcome was the ratio of the number of objects that had used 30/30 iterations divided by the number of objects that had used on 1 iteration at each 100-iteration interval. For example, at the 100^th^ iteration, the number of iterations that the object used during iterations 70 to 100 was calculated (i.e., the last “30 days”. As is clear from the figures (each generated using different starting seeds), the model is generally stable by the 1000^th^ iteration. Based on these results, we chose to use 2000 iterations to increase confidence in the robustness of the results.

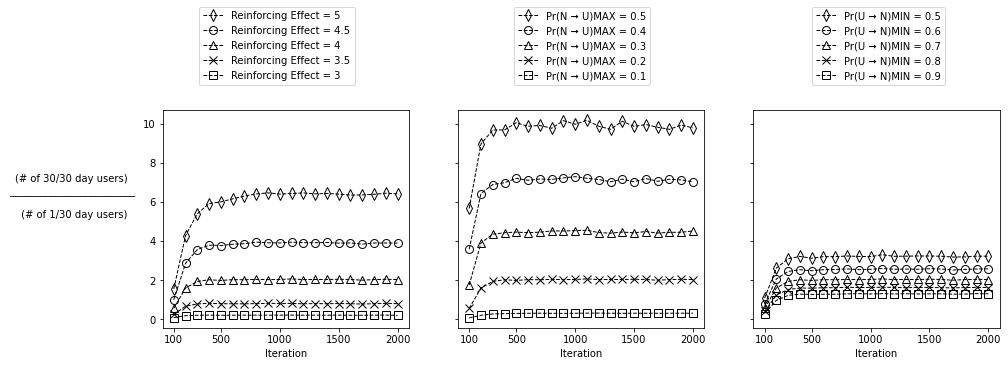

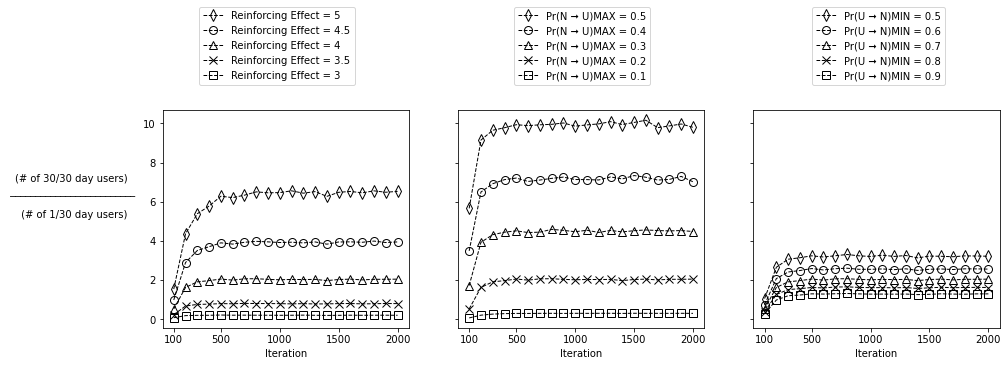

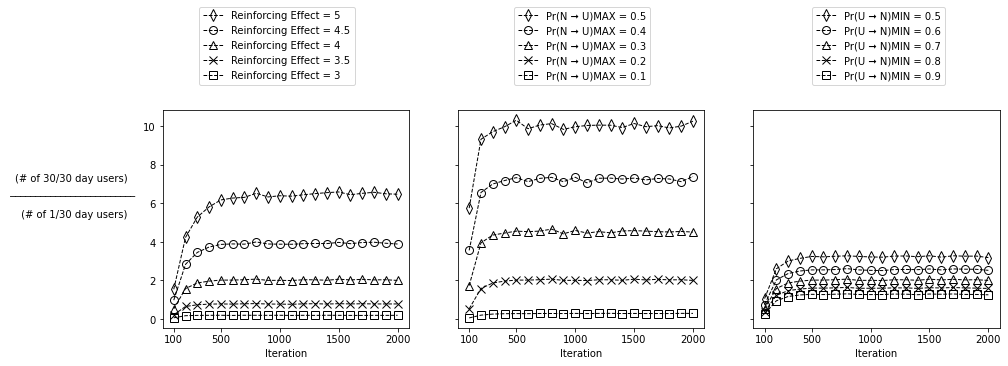

**Model Calibration**

For each substance (alcohol, cannabis, tobacco cigarettes), a least-squares approach was used to identify the best fit between a simulated distribution of past 30-day use and the empirical (NSDUH) distribution of past 3o-day use. Simulated distributions were generated using all possible combinations of values for three parameters: Maximum Risk Factors Effect, Minimum Protective Factors Effect, and Reinforcing Effect. Step 1 of this parameter space sweep tested values of the Maximum Risk Factors Effect ranging from 0.1 to 0.4 (0.05 denomination); values of the Minimum Protective Factors Effect ranging from 0.2 to 1 in (0.05 denomination); and values of the Reinforcing Effect ranging from 2.75 to 5.0 (0.25 denomination). For example, one simulation might examine the distribution produced when the Maximum Risk Factors Effect, Minimum Protective Factors Effect, and Reinforcing Effect parameters are set to values of 0.15, 0.75, and 3.25 respectively. A simulated distribution (using 2000 objects and 2000 iterations) was generated for each possible 3-value combination of parameter values and compared with the empirical distribution of a particular substance by calculating the sum of squared differences in the proportions associated with each possible past 30-day use frequency (e.g., simulated proportion that used on 5 days vs. empirical proportion that used on 5 days; 0 days of use was excluded). The combination of parameter values that produced the smallest sum of squared differences was considered optimal. In step 2, the entire process was repeated using a narrower range of parameter values with smaller denominations and 5000 computational objects. If the initial solution included a parameter value that was either the highest or lowest value of the range of values tested, then the simulation was conducted again using a wider range of parameter values. In step 3, the process was repeated a using n=10,000-20,000 computational objects. Multiple starting seeds were testing during the calibration process to avoid local minima.

**Testing non-uniform beta distributions**

In the primary study, the values of Pr(U→N)_i=0_ and Pr(N→U) were randomly drawn from uniform distributions. To examine the robustness of the results, we used non-uniform beta distributions and tested whether the model could still reproduce the “U-shaped” past 30-day use frequency distributions of interest in the study. We found that the model was indeed capable of reproducing these “U-shaped” distributions (right figure) using a non-uniform population distribution (left figure) for the initialization of the objects’ probabilities.

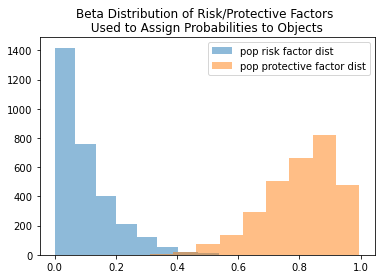

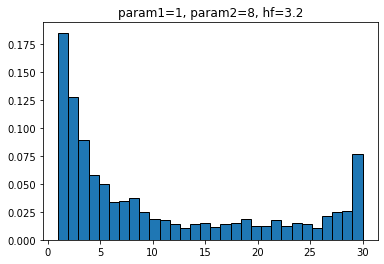

**Additional Material**

To understand how the updating process for Pr(U→N)_i>0_ works, consider the following example. If an object used the substance on 6 of the previous 30 iterations (i.e. mean of use history = 0.2), and the reinforcing effect parameter is set to a value of 1 by the modeler, then $e^{\left( -mean of use history * reinforcing effect \right)}$ is calculated as $e^{\left( -0.2 * 1 \right)}$ = 0.82. Multiplying Pr(U→N)_i=0_ by 0.82 yields the updated value of Pr(U→N)_i>0_. Stated differently, reducing Pr(U→N)_i=0_ by 18% yields the updated value of Pr(U→N)_i>0_ (Figure X). Thus, for example, if the fixed Pr(U→N)_i=0_ is 0.7, then the updated value for Pr(U→N)_i>0_ is ~0.57 (i.e., 0.7 * 0.82). This updated Pr(U→N)_i>0_ value (i.e., updated probability of discontinuing substance use) will be used by the object in the next iteration. Note that the value of Pr(U→N)_i=0_ does not change, meaning that the same value of Pr(U→N)_i=0_ is used at each iteration to calculate a new Pr(U→N)_i>0_.

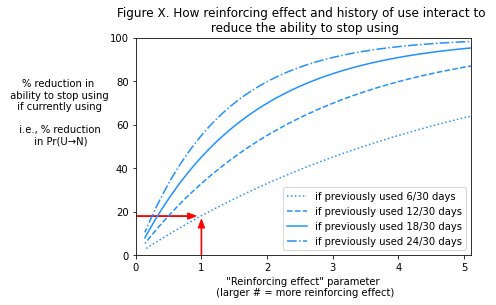
