## Supplementary material for "A simple computational model of population substance use": Model Code: figure2_code.py

```
# -*- coding: utf-8 -*-
"""
Created on Mon May 30 09:04:27 2022

@author: jacob
"""


import numpy as np
import random
import pandas as pd
import matplotlib.pyplot as plt
import seaborn as sns

########################################################################

class agent:
    def __init__(self, id):
      self.id = id
      self.p_tran_to_use = np.random.uniform(0, p_tran_to_use_max) # Pr(N U)
      self.p_tran_to_nouse_orig = np.random.uniform(p_tran_to_nouse_min, 1) # Pr(U N)
      self.p_tran_to_nouse_new = self.p_tran_to_nouse_orig
      self.eng = False 
      self.hist = [False] # EVERY AGENT STARTS OFF AS 0 SO THATS WHY HISTORY LENGTH WILL BE 1 LONGER THAN NUMBER OF ITERATIONS
      #self.p_tran_to_nouse_new_hist=[]      

    def engage(self):
        
      current_user = self.hist[-1]
      
      randomdraw = np.random.uniform(0,1)
        
      hist_mean = np.mean(self.hist[-hist_effect_length:]) 
      
      #YES PATH DEPENDENCE
      #habit_pressure = np.exp(-hist_mean * hab_factor) if len(self.hist) > hist_effect_length else 1 
      
      #NO PATH DEPENDENCE
      habit_pressure = 1
     
      #check if you will go back to NO use
      if current_user==True:
         self.eng = False if randomdraw < self.p_tran_to_nouse_new else True
                                                     #U_N              U_U
         """
         The above is a random draw based on your probability.
         If your p_tran_to_nouse threshold is 0.7 then there is a 70% chance that a random
         draw from a uniform dist will be less than the threshold and so there is
         a 70% that you will not use today if you HAD used yesterday
         """
      #if you are not a current user and your random draw is less than your prob of using, then you use 
      else: 
         self.eng = True if randomdraw < self.p_tran_to_use else False
                                             # N_U                N_N
                                             
      #If you used this iteration, re-calculate p_tran_to_nouse_new
      if self.eng == True:
         self.p_tran_to_nouse_new = self.p_tran_to_nouse_orig * habit_pressure # making the well deeper
         #habit pressure is number between 0 and 1

      
      self.hist.append(self.eng)
            
      #@@@@@@@@@@@@@@@@@@@@@@@@@@@@@@@@@@@@@@@@@@@@@@@@@@@@@@@@@@@@@@@@@@@@@@@@@@@@@@@@@@@@@@@@
      if len(self.hist) >= n_hist:  # if your history is longer than 30 days, used 30 days
          histeffective = n_hist 
      else: 
          histeffective = len(self.hist) # if you history is not longer than 30 days, then use your history length
          
      return sum(self.hist[-histeffective:]) 
      #@@@@@@@@@@@@@@@@@@@@@@@@@@@@@@@@@@@@@@@@@@@@@@@@@@@@@@@@@@@@@@@@@@@@@@@@@@@@@@@@@@@@@@@@

    def lempel_ziv (self, start_tick, stop_tick):
        string_ints = [str(int+0) for int in self.hist[start_tick:stop_tick]]
        str_of_ints = "". join(string_ints)
    
        keys_dict = {}
    
        ind = 0
        inc = 1
        while True:
            if not (len(str_of_ints) >= ind+inc):
                break
            sub_str = str_of_ints[ind:ind + inc]
            #print (sub_str,ind,inc)
            if sub_str in keys_dict:
                inc += 1
            else:
                keys_dict[sub_str] = 0
                ind += inc
                inc = 1
        return len(keys_dict) 


########################################################################

#COMPARE P30 DISTS AND MEAN COMPLEXITIES OF RESULTS WITH AND WITHOUT PATH DEPENDENCE EFFECTS
#IE EFFECT COMPARING:
      #habit_pressure = np.exp(-hist_mean * hab_factor) 
      #vs.
      #habit_pressure = 1 


########################################################################

# DO NOT CHANGE
n_hist = 30 # study window
hist_effect_length = 30 # num days that goes into impacting behavior
    
########################################################################

# MODELER CAN CHANGE THESE VALUES 
n_agents =1000 #
n_iter = 2000 

p_tran_to_use_max=0.2 #lower num shifts dist to the right
p_tran_to_nouse_min=0.7 # higher num shifts distribution to the left
hab_factor = 4

########################################################################

#Run simulation
np.random.seed(12345)
#np.random.seed(56789)
#np.random.seed(54321)

popul = [agent(i) for i in range(n_agents)]

for j in range(0, n_iter):
   x_data=[ag.engage() for ag in popul]
   if j%100==0:
     #plot_xn_hist(x_data)
     print(j)
   
     
filterpop=[]
for ag in popul:
   if any(ag.hist[-n_hist:])==True:
      filterpop.append(sum(ag.hist[-n_hist:]))
plt.hist(filterpop,bins=30, density=True)
plt.ylim(0,1)
plt.xlim(0,30)

days, counts = np.unique(filterpop, return_counts=True)
freqprop = counts/sum(counts)
print(sum(counts))
print(len(filterpop))
print(freqprop)
print(sum(freqprop))
print(len(freqprop))


df = pd.DataFrame()
df['days'] = days
df['freqprop'] = freqprop
#Yes Path Dependence
#df.to_excel('C:/Users/jacob/Box Sync/(1)_Dartmouth/(2)_Research/Theory paper/path_depend_effects_yesPD.xlsx')


#For no path dependence grp
freqprop2 = np.append(freqprop, np.zeros(17))
days2 = np.append(days, np.arange(14,31,1))
plt.bar(x=np.arange(1,31),height=freqprop2, color='blue',
         edgecolor="black", width=1)
plt.ylim(0,1)


df = pd.DataFrame()
df['days2'] = days2
df['freqprop2'] = freqprop2
#No Path Dependence
#df.to_excel('C:/Users/jacob/Box Sync/(1)_Dartmouth/(2)_Research/Theory paper/path_depend_effects_noPD.xlsx')


#############################################

#Comparison figure

#Laptop
df_yesPD = pd.read_excel("C:/Users/jacob/Box Sync/(1)_Dartmouth/(2)_Research/Theory paper/path_depend_effects_yesPD.xlsx")
df_noPD = pd.read_excel("C:/Users/jacob/Box Sync/(1)_Dartmouth/(2)_Research/Theory paper/path_depend_effects_noPD.xlsx")

#Desktop
df_yesPD = pd.read_excel("C:/Users/jacob/Box/(1)_Dartmouth/(2)_Research/Theory paper/path_depend_effects_yesPD.xlsx")
df_noPD = pd.read_excel("C:/Users/jacob/Box/(1)_Dartmouth/(2)_Research/Theory paper/path_depend_effects_noPD.xlsx")


fig, (ax1, ax2) = plt.subplots(1,2, sharey=True, figsize=(10, 5))
#fig, (ax1, ax2) = plt.subplots(2, sharex=True, figsize=(20, 20))
ax1.bar(x=np.arange(1,31,1), height=df_noPD['freqprop2'], color='dodgerblue', edgecolor="black", width=1  )
ax2.bar(x=np.arange(1,31,1), height=df_yesPD['freqprop'], color='dodgerblue', edgecolor="black", width=1  )
ax1.spines["right"].set_visible(False)
ax1.spines["top"].set_visible(False)
ax2.spines["right"].set_visible(False)
ax2.spines["top"].set_visible(False)
ax1.set_ylim(0,1)
ax2.set_ylim(0,1)
ax1.set_yticks([0,0.1,0.2,0.3,0.4,0.5,0.6,0.7,0.8,0.9,1.0])
ax1.set_yticklabels(['0','','20','','40','','60','','80','','100'],rotation=0,fontsize=15)

fig.suptitle('Figure 2. Simulated distributions of past 30-day substance use frequencies produced\nby disabling and enabling the path dependence function',
             x = -0.07, y=1.03,ha='left',fontsize=17)

xticklabels=['1',
             '5',
             '10',
             '15',
             '20',
             '25',
             '30',]
xticklocations = [1,5,10,15,20,25,30]

ax1.set_xticks(xticklocations)
ax2.set_xticks(xticklocations)
ax1.set_xticklabels(xticklabels,rotation=0,fontsize=15)
ax2.set_xticklabels(xticklabels,rotation=0,fontsize=15)

ax1.set_xlabel("Number days used \n in past 30 days", fontsize=17)
ax2.set_xlabel("Number days used \n in past 30 days", fontsize=17)
ax1.text(15,0.8, 'Path Dependence Function\nDisabled', fontsize=17,
                       bbox={'facecolor':'white','alpha':1,'edgecolor':'none','pad':1},
                       ha='center', va='center')
#ax1.text(15,0.5, f'Mean Lifetime\nLZ Complexity = {np.round(df_noPD.ave_lt_lzcomplex.mean(),2)} ', fontsize=17,
#                       bbox={'facecolor':'white','alpha':1,'edgecolor':'none','pad':1},
#                       ha='center', va='center')

ax2.text(15,0.8, 'Path Dependence Function\nEnabled', fontsize=17,
                       bbox={'facecolor':'white','alpha':1,'edgecolor':'none','pad':1},
                       ha='center', va='center')
#ax2.text(15,0.5, f'Mean Lifetime\nLZ Complexity = {np.round(df_yesPD.ave_lt_lzcomplex.mean(),2)} ', fontsize=17,
#                       bbox={'facecolor':'white','alpha':1,'edgecolor':'none','pad':1},
#                       ha='center', va='center')

ax1.set_ylabel("Percent of\npast-month\nconsumers", fontsize=15, rotation=0, labelpad=50)

ax1.text(-10,-0.4, 'Note: 100,000 objects and 2000 iterations used to created simulated distributions', fontsize=12,
                       bbox={'facecolor':'white','alpha':1,'edgecolor':'none','pad':1},
                       ha='left')

plt.subplots_adjust(left=0.1, 
                    bottom=None,
                    right=None,
                    top=None,
                    wspace=0.05,
                    hspace=0.1)


plt.savefig(r'C:\Users\jacob\Box Sync\(1)_Dartmouth\(2)_Research\Theory paper\Figure2.png', bbox_inches ="tight", format='png', pad_inches = 0.1, dpi=800,)
```
