## Supplementary material for "A simple computational model of population substance use": Model Code: figure3_code.py

```
# -*- coding: utf-8 -*-
"""
Created on Sat Apr 23 17:21:38 2022

@author: jacob
"""

import numpy as np
import random
import pandas as pd
import matplotlib.pyplot as plt
import seaborn as sns


#############################################
#############################################
#############################################
#############################################

#CREATE FIGURE

#These excel files were created in sim_dists_100k_2000_iter_v3.py

#Laptop
#Import 100k 2000 iteration simulated distributions
df_import_alc = pd.read_excel("C:/Users/jacob/Box Sync/(1)_Dartmouth/(2)_Research/Theory paper/alc_100kAG_2kITER_3_0165_05.xlsx")
df_import_mj = pd.read_excel("C:/Users/jacob/Box Sync/(1)_Dartmouth/(2)_Research/Theory paper/mj_100kAG_2kITER_38_0175_058.xlsx")
df_import_cig = pd.read_excel("C:/Users/jacob/Box Sync/(1)_Dartmouth/(2)_Research/Theory paper/cig_100kAG_2kITER_47_016_023.xlsx")

#Laptop
#Import cleaned NSDUH distributions
xls_p30alcdf_nsduh = pd.ExcelFile('C:/Users/jacob/Box Sync/(1)_Dartmouth/(2)_Research/Theory paper/nsduh_p30_alcfix.xlsx')
p30alcdf_nsduh = xls_p30alcdf_nsduh.parse(xls_p30alcdf_nsduh.sheet_names[0])
xls_p30mjdf_nsduh = pd.ExcelFile('C:/Users/jacob/Box Sync/(1)_Dartmouth/(2)_Research/Theory paper/nsduh_p30_mjfix.xlsx')
p30mjdf_nsduh = xls_p30mjdf_nsduh.parse(xls_p30mjdf_nsduh.sheet_names[0])
xls_p30cigdf_nsduh = pd.ExcelFile('C:/Users/jacob/Box Sync/(1)_Dartmouth/(2)_Research/Theory paper/nsduh_p30_cigfix.xlsx')
p30cigdf_nsduh = xls_p30cigdf_nsduh.parse(xls_p30cigdf_nsduh.sheet_names[0])

#Desktop
#Import 100k 2000 iteration simulated distributions
df_import_alc = pd.read_excel("C:/Users/jacob/Box/(1)_Dartmouth/(2)_Research/Theory paper/alc_100kAG_2kITER_3_0165_05.xlsx")
df_import_mj = pd.read_excel("C:/Users/jacob/Box/(1)_Dartmouth/(2)_Research/Theory paper/mj_100kAG_2kITER_38_0175_058.xlsx")
df_import_cig = pd.read_excel("C:/Users/jacob/Box/(1)_Dartmouth/(2)_Research/Theory paper/cig_100kAG_2kITER_47_016_023.xlsx")

#Desktop
#Import cleaned NSDUH distributions
xls_p30alcdf_nsduh = pd.ExcelFile('C:/Users/jacob/Box/(1)_Dartmouth/(2)_Research/Theory paper/nsduh_p30_alcfix.xlsx')
p30alcdf_nsduh = xls_p30alcdf_nsduh.parse(xls_p30alcdf_nsduh.sheet_names[0])
xls_p30mjdf_nsduh = pd.ExcelFile('C:/Users/jacob/Box/(1)_Dartmouth/(2)_Research/Theory paper/nsduh_p30_mjfix.xlsx')
p30mjdf_nsduh = xls_p30mjdf_nsduh.parse(xls_p30mjdf_nsduh.sheet_names[0])
xls_p30cigdf_nsduh = pd.ExcelFile('C:/Users/jacob/Box/(1)_Dartmouth/(2)_Research/Theory paper/nsduh_p30_cigfix.xlsx')
p30cigdf_nsduh = xls_p30cigdf_nsduh.parse(xls_p30cigdf_nsduh.sheet_names[0])

df_all = pd.DataFrame()
df_all['days'] = np.arange(1,31)
df_all['alc_sim_prop'] = df_import_alc['freqprop']
df_all['mj_sim_prop'] = df_import_mj['freqprop']
df_all['cig_sim_prop'] = df_import_cig['freqprop']
df_all['alc_nsduh_prop'] = p30alcdf_nsduh['props']
df_all['mj_nsduh_prop'] = p30mjdf_nsduh['props']
df_all['cig_nsduh_prop'] = p30cigdf_nsduh['props']
df_all['alc_nsduh_ci'] = p30alcdf_nsduh['ci']
df_all['mj_nsduh_ci'] = p30mjdf_nsduh['ci']
df_all['cig_nsduh_ci'] = p30cigdf_nsduh['ci']


alc_nsduh_prop=(0,0)
mj_nsduh_prop=(1,0)
cig_nsduh_prop=(2,0)

alc_sim_prop=(0,1)
mj_sim_prop=(1,1)
cig_sim_prop=(2,1)


fig, axs = plt.subplots(3, 2, sharex=True, figsize=(8, 11))
axs[alc_sim_prop].bar(df_all['days'],df_all['alc_sim_prop'], color='dodgerblue', edgecolor="black", width=1)
axs[mj_sim_prop].bar(df_all['days'],df_all['mj_sim_prop'], color='dodgerblue', edgecolor="black", width=1)
axs[cig_sim_prop].bar(df_all['days'],df_all['cig_sim_prop'], color='dodgerblue', edgecolor="black", width=1)
axs[alc_nsduh_prop].bar(df_all['days'],df_all['alc_nsduh_prop'], color='red', edgecolor="black", width=1)
axs[mj_nsduh_prop].bar(df_all['days'],df_all['mj_nsduh_prop'], color='red', edgecolor="black", width=1)
axs[cig_nsduh_prop].bar(df_all['days'],df_all['cig_nsduh_prop'], color='red', edgecolor="black", width=1)
axs[alc_sim_prop].spines["right"].set_visible(False)
axs[mj_sim_prop].spines["right"].set_visible(False)
axs[cig_sim_prop].spines["right"].set_visible(False)
axs[alc_nsduh_prop].spines["right"].set_visible(False)
axs[mj_nsduh_prop].spines["right"].set_visible(False)
axs[cig_nsduh_prop].spines["right"].set_visible(False)
axs[alc_sim_prop].spines["top"].set_visible(False)
axs[mj_sim_prop].spines["top"].set_visible(False)
axs[cig_sim_prop].spines["top"].set_visible(False)
axs[alc_nsduh_prop].spines["top"].set_visible(False)
axs[mj_nsduh_prop].spines["top"].set_visible(False)
axs[cig_nsduh_prop].spines["top"].set_visible(False)

axs[alc_sim_prop].set_ylim(0,1)
axs[mj_sim_prop].set_ylim(0,1)
axs[cig_sim_prop].set_ylim(0,1)
axs[alc_nsduh_prop].set_ylim(0,1)
axs[mj_nsduh_prop].set_ylim(0,1)
axs[cig_nsduh_prop].set_ylim(0,1)

axs[alc_sim_prop].set_yticks([0,0.1,0.2,0.3,0.4,0.5,0.6,0.7,0.8,0.9,1.0])
axs[mj_sim_prop].set_yticks([0,0.1,0.2,0.3,0.4,0.5,0.6,0.7,0.8,0.9,1.0])
axs[cig_sim_prop].set_yticks([0,0.1,0.2,0.3,0.4,0.5,0.6,0.7,0.8,0.9,1.0])
axs[alc_nsduh_prop].set_yticks([0,0.1,0.2,0.3,0.4,0.5,0.6,0.7,0.8,0.9,1.0])
axs[mj_nsduh_prop].set_yticks([0,0.1,0.2,0.3,0.4,0.5,0.6,0.7,0.8,0.9,1.0])
axs[cig_nsduh_prop].set_yticks([0,0.1,0.2,0.3,0.4,0.5,0.6,0.7,0.8,0.9,1.0])

axs[alc_nsduh_prop].set_yticklabels(['0','','20','','40','','60','','80','','100'],rotation=0,fontsize=15)
axs[mj_nsduh_prop].set_yticklabels(['0','','20','','40','','60','','80','','100'],rotation=0,fontsize=15)
axs[cig_nsduh_prop].set_yticklabels(['0','','20','','40','','60','','80','','100'],rotation=0,fontsize=15)

axs[alc_sim_prop].set_yticklabels(['','','','','','','','','','',''],rotation=0,fontsize=15)
axs[mj_sim_prop].set_yticklabels(['','','','','','','','','','',''],rotation=0,fontsize=15)
axs[cig_sim_prop].set_yticklabels(['','','','','','','','','','',''],rotation=0,fontsize=15)

xticklabels=['1',
             '5',
             '10',
             '15',
             '20',
             '25',
             '30',]
xticklocations = [1,5,10,15,20,25,30]

axs[alc_sim_prop].set_xticks(xticklocations)
axs[mj_sim_prop].set_xticks(xticklocations)
axs[cig_sim_prop].set_xticks(xticklocations)
axs[alc_nsduh_prop].set_xticks(xticklocations)
axs[mj_nsduh_prop].set_xticks(xticklocations)
axs[cig_nsduh_prop].set_xticks(xticklocations)
axs[alc_sim_prop].set_xticklabels(xticklabels,rotation=0,fontsize=15)
axs[mj_sim_prop].set_xticklabels(xticklabels,rotation=0,fontsize=15)
axs[cig_sim_prop].set_xticklabels(xticklabels,rotation=0,fontsize=15)
axs[alc_nsduh_prop].set_xticklabels(xticklabels,rotation=0,fontsize=15)
axs[mj_nsduh_prop].set_xticklabels(xticklabels,rotation=0,fontsize=15)
axs[cig_nsduh_prop].set_xticklabels(xticklabels,rotation=0,fontsize=15)

plt.subplots_adjust(left=0.1, 
                    bottom=None,
                    right=None,
                    top=None,
                    wspace=0.05,
                    hspace=0.1)


fig.suptitle('Figure 3. Survey-based distributions vs. simulated distributions of past 30-day\nsubstance use frequencies',
             fontsize=20, x=-0.1, y=1.01, ha='left')

axs[cig_sim_prop].set_xlabel("Number days used \n in past 30 days", fontsize=17)
axs[cig_nsduh_prop].set_xlabel("Number days used \n in past 30 days", fontsize=17)

axs[alc_sim_prop].text(15,1.2, 'Simulation', fontsize=17,
                       bbox={'facecolor':'white','alpha':1,'edgecolor':'none','pad':1},
                       ha='center', va='top')
axs[alc_nsduh_prop].text(15,1.2, 'National Survey on \nDrug Use and Health', fontsize=17,
                       bbox={'facecolor':'white','alpha':1,'edgecolor':'none','pad':1},
                       ha='center', va='top')
axs[alc_sim_prop].text(50,1.2, ('Simulation\nparameter values\nset by modeler'), fontsize=17,
                       bbox={'facecolor':'white','alpha':1,'edgecolor':'none','pad':1},
                       ha='center', va='top')

axs[alc_sim_prop].text(15,0.7, 'ALCOHOL', fontsize=17,
                       bbox={'facecolor':'white','alpha':1,'edgecolor':'none','pad':1},
                       ha='center', va='center')

axs[alc_nsduh_prop].text(15,0.7, 'ALCOHOL', fontsize=17,
                       bbox={'facecolor':'white','alpha':1,'edgecolor':'none','pad':1},
                       ha='center', va='center')

axs[mj_sim_prop].text(15,0.7, 'CANNABIS', fontsize=17,
                       bbox={'facecolor':'white','alpha':1,'edgecolor':'none','pad':1},
                       ha='center', va='center')


axs[mj_nsduh_prop].text(15,0.7, 'CANNABIS', fontsize=17,
                       bbox={'facecolor':'white','alpha':1,'edgecolor':'none','pad':1},
                       ha='center', va='center')

axs[cig_sim_prop].text(15,0.7, 'CIGARETTE', fontsize=17,
                       bbox={'facecolor':'white','alpha':1,'edgecolor':'none','pad':1},
                       ha='center', va='center')
axs[cig_nsduh_prop].text(15,0.7, 'CIGARETTE', fontsize=17,
                       bbox={'facecolor':'white','alpha':1,'edgecolor':'none','pad':1},
                       ha='center', va='center')

axs[mj_nsduh_prop].set_ylabel("Percent of\npast-month\nconsumers", fontsize=15, rotation=0, labelpad=50)

axs[cig_nsduh_prop].text(-10,-0.6, 'Note: 100,000 objects and 2000 iterations used to created simulated distributions', fontsize=12,
                       bbox={'facecolor':'white','alpha':1,'edgecolor':'none','pad':1},
                       ha='left')


reff_alc = '3.0'
reff_mj = '3.8'
reff_cig = '4.7'
#pnumax_string= u'Pr(N \u279E U)MAX = '
pnumax_string= 'Max Risk Effect = '
pnumax_alc = '0.165'
pnumax_mj = '0.175'
pnumax_cig = '0.160'
#punmin_string= u'Pr(U \u279E N)MIN = '
punmin_string= 'Min Protect Effect = '
punmin_alc = '0.50'
punmin_mj = '0.58'
punmin_cig = '0.23'


axs[alc_sim_prop].text(35,0.75, ('Reinforcing Effect = ' + 
                                reff_alc + '\n' +
                                pnumax_string + 
                                pnumax_alc + '\n' +
                                punmin_string + 
                                punmin_alc), fontsize=17,
                       bbox={'facecolor':'white','alpha':1,'edgecolor':'none','pad':1},
                       ha='left', va='top')


axs[mj_sim_prop].text(35,0.75, ('Reinforcing Effect = ' + 
                                reff_mj + '\n' +
                                pnumax_string + 
                                pnumax_mj + '\n' +
                                punmin_string + 
                                punmin_mj), fontsize=17,
                       bbox={'facecolor':'white','alpha':1,'edgecolor':'none','pad':1},
                       ha='left', va='top')

axs[cig_sim_prop].text(35,0.75, ('Reinforcing Effect = ' + 
                                reff_cig + '\n' +
                                pnumax_string + 
                                pnumax_cig + '\n' +
                                punmin_string + 
                                punmin_cig), fontsize=17,
                       bbox={'facecolor':'white','alpha':1,'edgecolor':'none','pad':1},
                       ha='left', va='top')

plt.savefig(r'C:\Users\jacob\Box Sync\(1)_Dartmouth\(2)_Research\Theory paper\Figure3.png', format='png', dpi=700,)


####################################################################
####################################################################
####################################################################
####################################################################
```
