## Supplementary material for "A simple computational model of population substance use": Model Code: Figure4_code.py

```
# -*- coding: utf-8 -*-
"""
Created on Fri Aug  6 17:31:58 2021

@author: jacob
"""

#Lifetime Complexity Intervals


import numpy as np
import random
import pandas as pd
import matplotlib.pyplot as plt
import seaborn as sns


########################################################################

class agent:
    def __init__(self, id):
      self.id = id
      self.p_tran_to_use = np.random.uniform(0, p_tran_to_use_max)
      self.p_tran_to_nouse_orig = np.random.uniform(p_tran_to_nouse_min, 1)
      self.p_tran_to_nouse_new = self.p_tran_to_nouse_orig
      self.eng = False 
      self.hist = [False]
      #self.p_tran_to_nouse_new_hist=[]      

    def engage(self):
        
      current_user=self.hist[-1]
      
      randomdraw = np.random.uniform(0,1)
        
      hist_mean = np.mean(self.hist[-hist_effect_length:]) 
      
      habit_pressure = np.exp(-hist_mean * hab_factor) if len(self.hist) > hist_effect_length else 1
         #habit pressure is number between 0 and 1
         #habit factor can be above or below 1
     
      #check if you will go back to NO use
      if current_user==True:
         self.eng = False if randomdraw < self.p_tran_to_nouse_new else True
         """
         The above is a random draw based on your probability.
         If your p_tran_to_nouse threshold is 0.7 then there is a 70% chance that a random
         draw from a uniform dist will be less than the threshold and so there is
         a 70% that you will not use today if you HAD used yesterday
         """
      #if you are not a current user and your random draw is less than your prob of using, then you use 
      else: 
         self.eng = True if randomdraw < self.p_tran_to_use else False
      
      if self.eng == True:
         self.p_tran_to_nouse_new = self.p_tran_to_nouse_orig * habit_pressure # making the well deeper
         #habit pressure is number between 0 and 1

      
      self.hist.append(self.eng)
      
      #self.p_tran_to_nouse_new_hist.append(self.p_tran_to_nouse_new) 
      
      #@@@@@@@@@@@@@@@@@@@@@@@@@@@@@@@@@@@@@@@@@@@@@@@@@@@@@@@@@@@@@@@@@@@@@@@@@@@@@@@@@@@@@@@@
      if len(self.hist) >= n_hist:  # if your history is longer than 30 days, used 30 days
          histeffective = n_hist 
      else: 
          histeffective = len(self.hist) # if you history is not longer than 30 days, then use your history length
          
      return sum(self.hist[-histeffective:]) 
      #@@@@@@@@@@@@@@@@@@@@@@@@@@@@@@@@@@@@@@@@@@@@@@@@@@@@@@@@@@@@@@@@@@@@@@@@@@@@@@@@@@@@@@@@


########################################################################

#GLOBAL FUNCTIONS

def lempel_ziv (listoflogicals):
    string_ints = [str(int+0) for int in listoflogicals]
    str_of_ints = "". join(string_ints)
    
    keys_dict = {}
    
    ind = 0
    inc = 1
    while True:
        if not (len(str_of_ints) >= ind+inc):
            break
        sub_str = str_of_ints[ind:ind + inc]
        #print (sub_str,ind,inc)
        if sub_str in keys_dict:
            inc += 1
        else:
            keys_dict[sub_str] = 0
            ind += inc
            inc = 1
    return len(keys_dict)

def complexity_chunking(lifetime_history, num_chunks):
    chunk_size = int(len(lifetime_history) / num_chunks) # should be 30
    lz_chunk_list=[]
    for chunk in range(num_chunks):
        lz_chunk = lempel_ziv(lifetime_history[chunk_size*chunk:chunk_size*(chunk+1)])
        lz_chunk_list.append(lz_chunk)
    return lz_chunk_list


def days_used_in_chunk(lifetime_history, num_chunks):
    chunk_size = int(len(lifetime_history) / num_chunks)
    days_chunk_list=[]
    for chunknum in range(num_chunks):
        num_days = sum(lifetime_history[chunk_size*chunknum:chunk_size*(chunknum+1)])
        days_chunk_list.append(num_days)
    return days_chunk_list


########################################################################

#Set parameters
n_agents = 3000 # Needs to be 100_000
n_iter = 900 # 30 * 30day chunks = 900 days
n_hist = 30
hist_effect_length = 30
p_tran_to_use_max=0.2 #lower num shifts dist to the right
p_tran_to_nouse_min=0.7 # higher num shifts distribution to the left
hab_factor = 5
number_chunks = int(n_iter/30)
#number_chunks = int(n_iter/50)
#number_chunks = int(n_iter/100)
print("\n number chunks OK") if n_iter%50==0 else print('\n number chunks NOT ok')

#*******************************************
#********************************************

#Is it possible to plot the average usage by ckunk? Also, median usage. 

#*******************************************
#*******************************************

#Run simulation
np.random.seed(12345)
#np.random.seed(54321)
popul = [agent(i) for i in range(n_agents)]

for j in range(n_iter):
   x_data=[ag.engage() for ag in popul]
   if j%100 ==0:
       print(j)
   
#Create dataset of results    
df_ag=pd.DataFrame(columns=['p_use','p_nouse_orig',
                            'lz_chunk_list', 'p30_at_last_tick',
                            'daysused_in_chunk', 'lt_prop_days_used', 
                            'lthist', 'daysusedfirst200','daysusedfirst400',
                            'daysused400_600','daysusedfirst700'])

irow=0
for ag in popul:
    print('row number = ',irow)
    ###########################################################
    #each agent gets a list of complexity scores for each 30-day chunk of their history
    df_ag.loc[irow,'lz_chunk_list'] = complexity_chunking(ag.hist, number_chunks) 
    ############################################################
    daysused_in_chunk = days_used_in_chunk(ag.hist, number_chunks) 
    daysusedp30=sum(ag.hist[-30:])
    lthist = ag.hist[:]
    daysusedfirst200 = sum(ag.hist[0:200])
    daysusedfirst400 = sum(ag.hist[0:400])
    daysused400_600 = sum(ag.hist[400:600])
    daysusedfirst700 = sum(ag.hist[0:700])
    propdaysusedlt = sum(ag.hist) / len(ag.hist)
    df_ag.loc[irow,'p_use'] = ag.p_tran_to_use
    df_ag.loc[irow,'p_nouse_orig'] = ag.p_tran_to_nouse_orig
    df_ag.loc[irow,'daysused_in_chunk'] = daysused_in_chunk
    #Use lifetime history to determine addiction/not addiction
    df_ag.loc[irow,'lt_prop_days_used'] = propdaysusedlt
    df_ag.loc[irow,'lthist'] = lthist


    irow+=1


#Export to Excel file for later use


df_ag2400iter_100kagents = df_ag.copy()
df_ag2400iter_100kagents.to_excel('C:/Users/jacob/Box Sync/(1)_Dartmouth/(2)_Research/Theory paper/lt_complex_chunk_2400iter_100kagents.xlsx')


#Laptop
df_ag = pd.read_excel('C:/Users/jacob/Box Sync/(1)_Dartmouth/(2)_Research/Theory paper/lt_complex_chunk_2400iter_100kagents.xlsx')

#Desktop
df_ag = pd.read_excel('C:/Users/jacob/Box/(1)_Dartmouth/(2)_Research/Theory paper/lt_complex_chunk_2400iter_100kagents.xlsx')


#Fix complexity chunk lists
from ast import literal_eval
df_ag['lzs'] = df_ag['lz_chunk_list'].apply(literal_eval)
df_ag['lzs'] = df_ag['lzs'].apply(lambda x: np.array(x)) # because axis = 1, the index of the series is the VARIABLES/COLUMNS (not the orignal row index values)


#######################################################################
#######################################################################
#######################################################################
#######################################################################

#PLOTS: MEAN COMPLEXITY FOR A GIVEN 30-DAY CHUNK GROUPED BY LIFETIME PROPORTION DAYS USED
from matplotlib.pyplot import figure

figure(figsize=(8, 6), dpi=500)
xticks=np.arange(0,15,1)
xticklblnums = np.arange(30,510,60)
xticklblstr = ['']*15
location=0
for i in range(0,len(xticklblnums)):
    print(i)
    print(location)
    if location<30:
        xticklblstr[location]=str(xticklblnums[i])
        location +=2

########## MEAN COMPLEXITY FOR ADDICTED ##########
#Use lifetime history to determine addiction/not addiction
df_ag_high = df_ag[(df_ag['lt_prop_days_used']>=0.7)]
complexity_scores2 = np.stack(df_ag_high['lzs'].values)
print(complexity_scores2[0:5,0:10])
print(df_ag_high['lzs'][0:5])
avg_complexity_score_chunks2 = np.mean(complexity_scores2,axis=0)
plt.plot(avg_complexity_score_chunks2[0:15], marker='o',
         mfc="None", mec='black', ms=7, linewidth=1, color='black')
#plt.ylim(7,11)
#plt.xlim(-1,29)
#plt.xticks(xticks, xticklblstr, rotation ='45', fontsize=10)

########## MEAN COMPLEXITY FOR EVERYONE ELSE IN MIDDLE ##########
#Use lifetime history to determine addiction/not addiction
df_ag_mid = df_ag[(df_ag['lt_prop_days_used']>0.2) & (df_ag['lt_prop_days_used']<0.7)]
complexity_scores3 = np.stack(df_ag_mid['lzs'].values)
avg_complexity_score_chunks3 =np.mean(complexity_scores3,axis=0)
plt.plot(avg_complexity_score_chunks3[0:15], marker='s',
         mfc="None", mec='black', ms=7, linewidth=1, color='black')
#plt.ylim(7,11)
#plt.xlim(-1,29)
#plt.xticks(xticks, xticklblstr, rotation ='45', fontsize=10)

########## MEAN COMPLEXITY FOR NON ADDICTED ##########
#Use lifetime history to determine addiction/not addiction
df_ag_low = df_ag[(df_ag['lt_prop_days_used']<=0.2)]
complexity_scores4 = np.stack(df_ag_low['lzs'].values)
avg_complexity_score_chunks4 =np.mean(complexity_scores4,axis=0)
plt.plot(avg_complexity_score_chunks4[0:15], marker='x',
         mfc="None", mec='black', ms=7, linewidth=1, color='black')
plt.ylim(7,11)
plt.xlim(-1,15)
plt.xticks(xticks, xticklblstr, rotation ='45', fontsize=10)
plt.xticks(xticks, xticklblstr, rotation ='45', fontsize=10)
plt.xlabel('Iteration Number')

plt.legend(['Lifetime high-frequency consumer*',
            'Lifetime moderate-frequency consumer*',
            'Lifetime low-frequency consumer*',],bbox_to_anchor=(0.25, 0.7), prop={'size': 10})
plt.ylabel('Mean \n past 30-iteration \ncomplexity', rotation=None, labelpad=40)
plt.title('Figure 4. Mean complexity of 30-iteration intervals stratified by \n proportion of total iterations used')
plt.text(y=5.6, x=-2, s='Notes: Simulation conducted using 100,000 objects and 2400 iterations\n*High-frequency = used on ≥70% of 2400 iterations\n*Moderate-frequency = used on >20% & <70% of 2400 iterations\n*Low-frequency = used on ≤ 20% of 2400 iterations ', fontsize=8)
plt.savefig(r'C:\Users\jacob\Box Sync\(1)_Dartmouth\(2)_Research\Theory paper\Figure4.png', format='png', dpi=500,)


#######################################################################
#######################################################################
#######################################################################
#######################################################################
#######################################################################
```
