## Supplementary material for "A simple computational model of population substance use": Model Code: Figure_5_code.py

```
# -*- coding: utf-8 -*-
"""
Created on Fri Apr 15 14:35:36 2022

@author: jacob
"""


import numpy as np
import pandas as pd
import matplotlib.pyplot as plt


########################################################################

class agent:
    def __init__(self, id):
        self.id = id
        self.p_tran_to_use = np.random.uniform(0, p_tran_to_use_max)
        self.p_tran_to_nouse_orig = np.random.uniform(p_tran_to_nouse_min, 1)
        self.p_tran_to_nouse_new = self.p_tran_to_nouse_orig
        self.eng = False
        # EVERY AGENT STARTS OFF AS 0 SO THATS WHY HISTORY LENGTH WILL BE 1 LONGER THAN NUMBER OF ITERATIONS
        self.hist = [False]
        # self.p_tran_to_nouse_new_hist=[]

    def engage(self):

        current_user = self.hist[-1]

        randomdraw = np.random.uniform(0, 1)

        #hist_mean = np.mean(self.hist[-n_hist:])
        # USING A LOWER NUMBER HERE CAUSES THE SAME THING AS USING A LOWER H50 NUMBER IN THE MAIN SECTION
        hist_mean = np.mean(self.hist[-hist_effect_length:])

        # removing if statement seems to still work and not give 30-day bump
        habit_pressure = np.exp(-hist_mean *
                                hab_factor) if len(self.hist) > hist_effect_length else 1
        # habit pressure is number between 0 and 1
        # habit factor can be above or below 1

        # check if you will go back to NO use
        if current_user == True:
            self.eng = False if randomdraw < self.p_tran_to_nouse_new else True
            # U_N              U_U
            """
         The above is a random draw based on your probability.
         If your p_tran_to_nouse threshold is 0.7 then there is a 70% chance that a random
         draw from a uniform dist will be less than the threshold and so there is
         a 70% that you will not use today if you HAD used yesterday
         """
        # if you are not a current user and your random draw is less than your prob of using, then you use
        else:
            self.eng = True if randomdraw < self.p_tran_to_use else False
            # N_U                N_N

        # If you used this iteration, re-calculate p_tran_to_nouse_new
        if self.eng == True:
            self.p_tran_to_nouse_new = self.p_tran_to_nouse_orig * \
                habit_pressure  # making the well deeper
            # habit pressure is number between 0 and 1

        self.hist.append(self.eng)

        # self.p_tran_to_nouse_new_hist.append(self.p_tran_to_nouse_new)

        # @@@@@@@@@@@@@@@@@@@@@@@@@@@@@@@@@@@@@@@@@@@@@@@@@@@@@@@@@@@@@@@@@@@@@@@@@@@@@@@@@@@@@@@@
        if len(self.hist) >= n_hist:  # if your history is longer than 30 days, used 30 days
            histeffective = n_hist
        else:
            # if you history is not longer than 30 days, then use your history length
            histeffective = len(self.hist)

        return sum(self.hist[-histeffective:])
        # @@@@@@@@@@@@@@@@@@@@@@@@@@@@@@@@@@@@@@@@@@@@@@@@@@@@@@@@@@@@@@@@@@@@@@@@@@@@@@@@@@@@@@@@


def lempel_ziv(listoflogicals):
    string_ints = [str(int+0) for int in listoflogicals]
    str_of_ints = "". join(string_ints)

    keys_dict = {}

    ind = 0
    inc = 1
    while True:
        if not (len(str_of_ints) >= ind+inc):
            break
        sub_str = str_of_ints[ind:ind + inc]
        #print (sub_str,ind,inc)
        if sub_str in keys_dict:
            inc += 1
        else:
            keys_dict[sub_str] = 0
            ind += inc
            inc = 1
    return len(keys_dict)


########################################################################

# Set parameters (Alcohol) based on OLS optim results
n_agents = 100_000
# at start self.hist = [False] which means EVERY AGENT STARTS OFF AS 0 SO THATS WHY HISTORY LENGTH WILL BE 1 LONGER THAN NUMBER OF ITERATIONS
n_iter = 2000
n_hist = 30  # study window
hist_effect_length = 30
p_tran_to_use_max = 0.165
p_tran_to_nouse_min = 0.5
hab_factor = 3

########################################################################

# Run simulation
np.random.seed(12345)

popul = [agent(i) for i in range(n_agents)]

for j in range(0, n_iter):
    x_data = [ag.engage() for ag in popul]
    if j % 100 == 0:
        # plot_xn_hist(x_data)
        print(j)

for ag in popul:
    print(ag.id, sum(ag.hist))

sum(popul[28].hist)

df_ag = pd.DataFrame(columns=['p_use', 'p_nouse_orig',
                              'lt_prop_days_used',
                              'lthist', ])

irow = 0
for ag in popul:
    print('row number = ', irow)
    lthist = ag.hist[:]
    propdaysusedlt = sum(ag.hist) / len(ag.hist)
    df_ag.loc[irow, 'p_use'] = ag.p_tran_to_use
    df_ag.loc[irow, 'p_nouse_orig'] = ag.p_tran_to_nouse_orig
    df_ag.loc[irow, 'lt_prop_days_used'] = propdaysusedlt
    df_ag.loc[irow, 'lthist'] = lthist

    irow += 1


# EXPORT DATA to Excel file for later use

df_streak_alc = df_ag.copy()
# LAPTOP
df_streak_alc.to_excel(
    'C:/Users/jacob/Box Sync/(1)_Dartmouth/(2)_Research/Theory paper/df_streak_alc_100k_2000iter.xlsx')
# DESKTOP
df_streak_alc.to_excel(
    'C:/Users/jacob/Box/(1)_Dartmouth/(2)_Research/Theory paper/df_streak_alc_100k_2000iter.xlsx')


######################################################################


# IMPORT DATA FOR FIGURE
# LAPTOP
df_streak = pd.read_excel(
    'C:/Users/jacob/Box Sync/(1)_Dartmouth/(2)_Research/Theory paper/df_streak_alc_100k_2000iter.xlsx')
# DESKTOP
df_streak = pd.read_excel(
    'C:/Users/jacob/Box/(1)_Dartmouth/(2)_Research/Theory paper/df_streak_alc_100k_2000iter.xlsx')

# -----------------------------------------------
# Use this to identify objects with different combos of high/low complexity & high/low proportion days used


def propdays_and_complexity(rownum, dataframe):
    myary = dataframe.loc[rownum, 'lthist'].strip('][').split(', ')
    myary2 = [True if x == 'True' else False for x in myary]
    myary2_props = sum(myary2)/len(myary2)
    myary2_lz = lempel_ziv(myary2)
    return np.round(myary2_props, 3), myary2_lz
    #print('proportion = ',np.round(myary2_props,3),'\n','lz = ',myary2_lz)


# Find objects that are good examples of diff proportion/complexity relations
highlz_ids = []
for i in range(0, 100_000):
    prop, lz = propdays_and_complexity(i, df_streak)
    if 0.79 < prop < 0.86:
        print(i)
        # print('prop=',prop)
        # print('lz=',lz)
        # print('---------------')
        # print('\n')
        highlz_ids.append([i, lz])

highlz = [pair for pair in highlz_ids if pair[1] > 140]
for i in highlz:
    print(i)
    print(propdays_and_complexity(i[0], df_streak))
    print('------------')
    print('\n')
print(propdays_and_complexity(62422, df_streak))
print(propdays_and_complexity(22439, df_streak))


lowlz_ids = []
for i in range(0, 100_000):
    prop, lz = propdays_and_complexity(i, df_streak)
    if 0.054 < prop < 0.057:
        print(i)
        # print('prop=',prop)
        # print('lz=',lz)
        # print('---------------')
        # print('\n')
        lowlz_ids.append([i, lz])

lowlz = [pair for pair in lowlz_ids if pair[1] < 140]
print(len(lowlz))
for i in lowlz[:250]:
    print(i)
    print(propdays_and_complexity(i[0], df_streak))
    print('------------')
    print('\n')
print(propdays_and_complexity(39655, df_streak))
print(propdays_and_complexity(39305, df_streak))


def streak(ary, suppress0=True):
    ary = np.array(ary)*1
    if not suppress0:
        ary[ary == 0] = -1
    pos = np.clip(ary, 0, 1).astype(bool).cumsum()
    neg = np.clip(ary, -1, 0).astype(bool).cumsum()
    return np.where(ary >= 0, pos-np.maximum.accumulate(np.where(ary <= 0, pos, 0)),
                    -neg+np.maximum.accumulate(np.where(ary >= 0, neg, 0)))


# -----------------------------------
# Low use/Low Complexity
myarylowlow = df_streak.loc[39655, 'lthist'].strip('][').split(', ')
print(myarylowlow)
myarylowlow2 = [True if x == 'True' else False for x in myarylowlow]
print(myarylowlow2)
myarylowlow2_prop = sum(myarylowlow2)/len(myarylowlow2)
myarylowlow2_lz = lempel_ziv(myarylowlow2)
mystreaklowlow = streak(myarylowlow2, suppress0=False)
print(mystreaklowlow)
streak_pos_lowlow = mystreaklowlow.copy()
streak_pos_lowlow[streak_pos_lowlow < 0] = 0
streak_neg_lowlow = mystreaklowlow.copy()
streak_neg_lowlow[streak_neg_lowlow > 0] = 0
streak_xaxis_lowlow = np.arange(len(mystreaklowlow))
#plt.bar(streak_xaxis_low, streak_pos_low, color='red', edgecolor='red')
#plt.bar(streak_xaxis_low, streak_neg_low, color='green',  edgecolor='green')
# -----------------------------------
# Low use/High Complexity
myarylowhigh = df_streak.loc[39305, 'lthist'].strip('][').split(', ')
print(myarylowhigh)
myarylowhigh2 = [True if x == 'True' else False for x in myarylowhigh]
print(myarylowhigh2)
myarylowhigh2_prop = sum(myarylowhigh2)/len(myarylowhigh2)
myarylowhigh2_lz = lempel_ziv(myarylowhigh2)
mystreaklowhigh = streak(myarylowhigh2, suppress0=False)
print(mystreaklowhigh)
streak_pos_lowhigh = mystreaklowhigh.copy()
streak_pos_lowhigh[streak_pos_lowhigh < 0] = 0
streak_neg_lowhigh = mystreaklowhigh.copy()
streak_neg_lowhigh[streak_neg_lowhigh > 0] = 0
streak_xaxis_lowhigh = np.arange(len(mystreaklowhigh))
#plt.bar(streak_xaxis_low, streak_pos_low, color='red', edgecolor='red')
#plt.bar(streak_xaxis_low, streak_neg_low, color='green',  edgecolor='green')
# -----------------------------------
# High use/Low Complexity
myaryhighlow = df_streak.loc[22439, 'lthist'].strip('][').split(', ')
print(myaryhighlow)
myaryhighlow2 = [True if x == 'True' else False for x in myaryhighlow]
print(myaryhighlow2)
myaryhighlow2_prop = sum(myaryhighlow2)/len(myaryhighlow2)
myaryhighlow2_lz = lempel_ziv(myaryhighlow2)
mystreakhighlow = streak(myaryhighlow2, suppress0=False)
print(mystreakhighlow)
streak_pos_highlow = mystreakhighlow.copy()
streak_pos_highlow[streak_pos_highlow < 0] = 0
streak_neg_highlow = mystreakhighlow.copy()
streak_neg_highlow[streak_neg_highlow > 0] = 0
streak_xaxis_highlow = np.arange(len(mystreakhighlow))
#plt.bar(streak_xaxis_highlow, streak_pos_highlow, color='red', edgecolor='red')
#plt.bar(streak_xaxis_highlow, streak_neg_highlow, color='green',  edgecolor='green')
# -----------------------------------
# High use/High Complexity
myaryhighhigh = df_streak.loc[62422, 'lthist'].strip('][').split(', ')
print(myaryhighhigh)
myaryhighhigh2 = [True if x == 'True' else False for x in myaryhighhigh]
print(myaryhighhigh2)
myaryhighhigh2_prop = sum(myaryhighhigh2)/len(myaryhighhigh2)
myaryhighhigh2_lz = lempel_ziv(myaryhighhigh2)
mystreakhighhigh = streak(myaryhighhigh2, suppress0=False)
print(mystreakhighhigh)
streak_pos_highhigh = mystreakhighhigh.copy()
streak_pos_highhigh[streak_pos_highhigh < 0] = 0
streak_neg_highhigh = mystreakhighhigh.copy()
streak_neg_highhigh[streak_neg_highhigh > 0] = 0
streak_xaxis_highhigh = np.arange(len(mystreakhighhigh))
#plt.bar(streak_xaxis_highhigh, streak_pos_highhigh, color='red', edgecolor='red')
#plt.bar(streak_xaxis_highhigh, streak_neg_highhigh, color='green',  edgecolor='green')


# Create Legend subgraph
legendlen = 50
legendx = np.arange(0, legendlen, 1)
legend_pos_streak = np.append(np.arange(0, int(legendlen/2), 1),
                              np.zeros(int(legendlen/2)))
legend_neg_streak = np.append(np.zeros(int(legendlen/2)),
                              np.arange(0, int(-1*legendlen/2), -1))


fig, axs = plt.subplots(2, 2, figsize=(12, 6))
axs[(0, 0)].bar(streak_xaxis_highlow, streak_pos_highlow,
                color='goldenrod', edgecolor='goldenrod')
axs[(0, 0)].bar(streak_xaxis_highlow,
                streak_neg_highlow, color='gray',  edgecolor='gray')
axs[(0, 1)].bar(streak_xaxis_highhigh, streak_pos_highhigh,
                color='goldenrod', edgecolor='goldenrod')
axs[(0, 1)].bar(streak_xaxis_highhigh,
                streak_neg_highhigh, color='gray',  edgecolor='gray')
axs[(1, 0)].bar(streak_xaxis_lowlow, streak_pos_lowlow,
                color='goldenrod', edgecolor='goldenrod')
axs[(1, 0)].bar(streak_xaxis_lowlow, streak_neg_lowlow,
                color='gray',  edgecolor='gray')
axs[(1, 1)].bar(streak_xaxis_lowhigh, streak_pos_lowhigh,
                color='goldenrod', edgecolor='goldenrod')
axs[(1, 1)].bar(streak_xaxis_lowhigh,
                streak_neg_lowhigh, color='gray',  edgecolor='gray')

left, bottom, width, height = [0.48, 0.75, 0.08, 0.12]
ax2 = fig.add_axes([left, bottom, width, height])
ax2.bar(legendx, legend_pos_streak, color='goldenrod', width=1)
ax2.bar(legendx, legend_neg_streak, color='gray', width=1)
# ax2.spines["right"].set_visible(False)
# ax2.spines["top"].set_visible(False)
# ax2.spines["bottom"].set_visible(False)
# ax2.spines["left"].set_visible(False)
# ax2.spines["left"].set_visible(False)
# ax2.axis('off')
ax2.set_xticks([])
ax2.set_yticks([])
redtext = left+0.1
greentext = redtext+0.044
fig.text(redtext-0.015, 0.8, '= ', fontsize=12, color='black')
fig.text(redtext, 0.8, '11111', fontsize=12, color='goldenrod')
fig.text(greentext, 0.8, '00000', fontsize=12, color='gray')
fig.text(0.5, 0.89, 'Legend', fontsize=12, color='black')


axs[(0, 0)].set_ylim(-50, 320)
axs[(0, 1)].set_ylim(-50, 320)
axs[(1, 0)].set_ylim(-600, max(streak_pos_lowlow))
axs[(1, 1)].set_ylim(-600, max(streak_pos_lowlow))

axs[(1, 0)].set_xlabel('Iteration Number')
axs[(1, 1)].set_xlabel('Iteration Number')

# ax2.set_ylim(-50,150)
#plt.xlabel("Iteration number", fontsize=13)
axs[(0, 0)].spines["right"].set_visible(False)
axs[(0, 0)].spines["top"].set_visible(False)
axs[(0, 0)].spines["bottom"].set_visible(False)
axs[(1, 0)].spines["right"].set_visible(False)
axs[(1, 0)].spines["top"].set_visible(False)
axs[(0, 1)].spines["right"].set_visible(False)
axs[(0, 1)].spines["top"].set_visible(False)
axs[(0, 1)].spines["left"].set_visible(False)
axs[(0, 1)].spines["bottom"].set_visible(False)
axs[(1, 1)].spines["right"].set_visible(False)
axs[(1, 1)].spines["top"].set_visible(False)
axs[(1, 1)].spines["left"].set_visible(False)

axs[(0, 0)].set_xticks([])
axs[(1, 1)].set_yticks([])
axs[(0, 1)].set_xticks([])
axs[(0, 1)].set_yticks([])


textfontsize = 9
axs[(0, 0)].text(0, 300, "Computational Object # 1", fontsize=textfontsize,
                 bbox={'facecolor': 'white', 'alpha': 1,
                       'edgecolor': 'none', 'pad': 1},
                 ha='left', va='top')
axs[(0, 0)].text(0, 250, f'Complexity = {myaryhighlow2_lz}', fontsize=textfontsize,
                 bbox={'facecolor': 'white', 'alpha': 1,
                       'edgecolor': 'none', 'pad': 1},
                 ha='left', va='top')
axs[(0, 0)].text(0, 200, f'Used {np.round(myaryhighlow2_prop*100,1)}% of iterations', fontsize=textfontsize,
                 bbox={'facecolor': 'white', 'alpha': 1,
                       'edgecolor': 'none', 'pad': 1},
                 ha='left', va='top')
axs[(0, 0)].text(1000, 275, "Spike represents\n~300 1's in a row", fontsize=textfontsize,
                 bbox={'facecolor': 'white', 'alpha': 1,
                       'edgecolor': 'none', 'pad': 1},
                 ha='left', va='top')
axs[(0, 0)].text(1650, 275, u"\u279E", fontsize=18,
                 bbox={'facecolor': 'white', 'alpha': 1,
                       'edgecolor': 'none', 'pad': 1},
                 ha='left', va='top')

axs[(0, 1)].text(1200, 300, "Computational Object # 2", fontsize=textfontsize,
                 bbox={'facecolor': 'white', 'alpha': 1,
                       'edgecolor': 'none', 'pad': 1},
                 ha='left', va='top')
axs[(0, 1)].text(1200, 250, f'Complexity = {myaryhighhigh2_lz}', fontsize=textfontsize,
                 bbox={'facecolor': 'white', 'alpha': 1,
                       'edgecolor': 'none', 'pad': 1},
                 ha='left', va='top')
axs[(0, 1)].text(1200, 200, f'Used {np.round(myaryhighhigh2_prop*100,1)}% of iterations', fontsize=textfontsize,
                 bbox={'facecolor': 'white', 'alpha': 1,
                       'edgecolor': 'none', 'pad': 1},
                 ha='left', va='top')

axs[(1, 0)].text(0, -300, "Computational Object # 3", fontsize=textfontsize,
                 bbox={'facecolor': 'white', 'alpha': 1,
                       'edgecolor': 'none', 'pad': 1},
                 ha='left', va='top')
axs[(1, 0)].text(0, -400, f'Complexity = {myarylowlow2_lz}', fontsize=textfontsize,
                 bbox={'facecolor': 'white', 'alpha': 1,
                       'edgecolor': 'none', 'pad': 1},
                 ha='left', va='top')
axs[(1, 0)].text(0, -500, f'Used {np.round(myarylowlow2_prop*100,1)}% of iterations', fontsize=textfontsize,
                 bbox={'facecolor': 'white', 'alpha': 1,
                       'edgecolor': 'none', 'pad': 1},
                 ha='left', va='top')
axs[(1, 0)].text(1350, -375, "Spike\nrepresents\n~500 0's\nin a row", fontsize=textfontsize,
                 bbox={'facecolor': 'white', 'alpha': 1,
                       'edgecolor': 'none', 'pad': 1},
                 ha='left', va='top')
axs[(1, 0)].text(1750, -400, u"\u279E", fontsize=18,
                 bbox={'facecolor': 'white', 'alpha': 1,
                       'edgecolor': 'none', 'pad': 1},
                 ha='left', va='top')

axs[(1, 1)].text(0, -300, "Computational Object # 4", fontsize=textfontsize,
                 bbox={'facecolor': 'white', 'alpha': 1,
                       'edgecolor': 'none', 'pad': 1},
                 ha='left', va='top')
axs[(1, 1)].text(0, -400, f'Complexity = {myarylowhigh2_lz}', fontsize=textfontsize,
                 bbox={'facecolor': 'white', 'alpha': 1,
                       'edgecolor': 'none', 'pad': 1},
                 ha='left', va='top')
axs[(1, 1)].text(0, -500, f'Used {np.round(myarylowhigh2_prop*100,1)}% of iterations', fontsize=textfontsize,
                 bbox={'facecolor': 'white', 'alpha': 1,
                       'edgecolor': 'none', 'pad': 1},
                 ha='left', va='top')

fig.suptitle("Figure 5. Pattern of Using (1's) and Not Using (0's) streaks over 2000 iterations for four different computational objects")
axs[(0, 0)].set_ylabel("Streak of use/no use", fontsize=13, labelpad=1)
axs[(1, 0)].set_ylabel("Streak of use/no use", fontsize=13, labelpad=1)
```
