## Supplementary material for "A simple computational model of population substance use": Model Code: non_uniform_dist_test.py

```
# Demonstration that non-uniform population distribution used to obtain risk and protective probabilities will still produce the U-shaped distribution of p30 days used.

"""
#Feel free to email me if you have any questions or
want to set up a Zoom call to go through the model



"""

import numpy as np
import matplotlib.pyplot as plt

########################################################################

#DO NOT CHANGE

class agent:
    def __init__(self, id):
      self.id = id
      #---------------------------------------------------------
      # BETA DISTRIBUTION INSTEAD OF UNIFORM DIST
      self.p_tran_to_use = np.random.beta(param1, 10-param1)   #risk factors
      self.p_tran_to_nouse_orig = np.random.beta(param2, 10-param2)   #protective factors
      #---------------------------------------------------------
      self.p_tran_to_nouse_new = self.p_tran_to_nouse_orig
      self.eng = False 
      self.hist = [False]      

    def engage(self):
        
      current_user = self.hist[-1]
      
      randomdraw = np.random.uniform(0,1)
        
      hist_mean = np.mean(self.hist[-hist_effect_length:]) 
      
      habit_pressure = np.exp(-hist_mean * hab_factor) if len(self.hist) > hist_effect_length else 1 
     
      #check if you will go back to NO use
      if current_user==True:
         self.eng = False if randomdraw < self.p_tran_to_nouse_new else True
                                                     #U_N              U_U
         """
         The above is a random draw based on your probability.
         If your p_tran_to_nouse threshold is 0.7 then there is a 70% chance that a random
         draw from a uniform dist will be less than the threshold and so there is
         a 70% that you will not use today if you HAD used yesterday
         """
      #if you are not a current user and your random draw is less than your prob of using, then you use 
      else: 
         self.eng = True if randomdraw < self.p_tran_to_use else False
                                             # N_U                N_N
                                             
      #If you used this iteration, re-calculate p_tran_to_nouse_new
      if self.eng == True:
         self.p_tran_to_nouse_new = self.p_tran_to_nouse_orig * habit_pressure 
         #habit pressure is number between 0 and 1

      
      self.hist.append(self.eng)
      
     
      #@@@@@@@@@@@@@@@@@@@@@@@@@@@@@@@@@@@@@@@@@@@@@@@@@@@@@@@@@@@@@@@@@@@@@@@@@@@@@@@@@@@@@@@@
      if len(self.hist) >= n_hist:  # if your history is longer than 30 days, used 30 days
          histeffective = n_hist 
      else: 
          histeffective = len(self.hist) # if you history is not longer than 30 days, then use your history length
          
      return sum(self.hist[-histeffective:]) 
      #@@@@@@@@@@@@@@@@@@@@@@@@@@@@@@@@@@@@@@@@@@@@@@@@@@@@@@@@@@@@@@@@@@@@@@@@@@@@@@@@@@@@@@@@

    def lempel_ziv (self, start_tick, stop_tick):
        string_ints = [str(int+0) for int in self.hist[start_tick:stop_tick]]
        str_of_ints = "". join(string_ints)
    
        keys_dict = {}
    
        ind = 0
        inc = 1
        while True:
            if not (len(str_of_ints) >= ind+inc):
                break
            sub_str = str_of_ints[ind:ind + inc]
            #print (sub_str,ind,inc)
            if sub_str in keys_dict:
                inc += 1
            else:
                keys_dict[sub_str] = 0
                ind += inc
                inc = 1
        return len(keys_dict) 


########################################################################

# DO NOT CHANGE
n_hist = 30 # study window
hist_effect_length = 30 # num days that goes into impacting behavior
    


########################################################################

# MODELER CAN CHANGE THESE VALUES 
n_agents =3000 # number of agents
n_iter = 2000 # number of iterations

param1 = 1 # risk
param2 = 8 # protective
hab_factor = 3.2


#--------------------------------------------------
#plt.hist(np.random.beta(param1, 10-param1,3000),alpha=0.5)  #risk
#plt.hist(np.random.beta(param2, 10-param2,3000),alpha=0.5)  #protect
#plt.legend(['pop risk factor dist', 'pop protective factor dist'])
#plt.title('Beta Distribution of Risk/Protective Factors \n Used to Assign Probabilities to Objects ')
#--------------------------------------------------

########################################################################

#Run simulation
np.random.seed(12345) 
"""
# MAKE SURE SEED IS THE SAME FOR ALL SCRIPTS
"""
popul = [agent(i) for i in range(n_agents)]

for j in range(0, n_iter):
   x_data=[ag.engage() for ag in popul]
   if j%100==0:
     #plot_xn_hist(x_data)
     print(j)
   
########################################################################

#Plot results/distribution  (removing those with 0/30 days of use)   

#Hisotgram
filterpop=[]
for ag in popul:
   if any(ag.hist[-n_hist:])==True:
      filterpop.append(sum(ag.hist[-n_hist:]))
plt.hist(filterpop,bins=30, density=True, edgecolor='black')
plt.title(f'param1={param1}, param2={param2}, hf={hab_factor}')

########################################################################
```
